# Including gender-specific features in epidemic modeling: the case of the second wave of COVID-19 in Italy

**DOI:** 10.64898/2025.12.19.25342675

**Authors:** Alessandro De Gaetano, Pietro Coletti, Nicola Perra, Alain Barrat, Daniela Paolotti

## Abstract

Biological and behavioral differences between genders influence infectious disease dynamics. Yet, most epidemiological models overlook these aspects in favor of age stratification alone. Here, we systematically evaluate the impact of incorporating gender-specific features into an age-structured epidemic compartmental model, calibrated to COVID-19 mortality data from the second wave in Italy (Autumn 2020–Winter 2021). We develop eight model versions representing different combinations of three data-driven features: gender-stratified contact matrices derived from CoMix data, gender-specific infection fatality ratios (IFR), and gender-dependent transmission rates linked to behavioral differences. We calibrate these models against aggregated mortality data and evaluate their performance on data disaggregated by gender, age, and both. Our results demonstrate that models incorporating gender-stratified contact patterns significantly outperform those relying solely on age, improving the accuracy of the fit even when analyzing age-disaggregated data alone. Furthermore, the inclusion of gender-specific IFR is essential for reproducing the empirically higher mortality rates observed in males. While phenomenological behavioral adjustments improve the fit for specific subgroups, such as older males, we observe trade-offs where maximizing performance for one demographic group occasionally reduces accuracy for another. Overall, our findings highlight that integrating gender data—particularly regarding contact patterns—is a critical step toward increasing the realism and precision of epidemiological models, even when outcome data is not fully disaggregated.

**Author summary:** Due to a combination of behavioral and biological factors, such as the adoption of protective measures and immune responses, infectious diseases affect men and women differently. However, the mathematical models used to understand and predict epidemics often overlook these aspects, focusing primarily on age. Here, we investigate whether explicitly including gender-specific data improves the accuracy of epidemic simulations. Using data from the second wave of the COVID-19 pandemic in Italy, we develop and compare several models that incorporated gender differences in social interaction patterns, biological risk of death, and protective behaviors. We find that men and women exhibit distinct social mixing patterns, and including this information is the most important factor for accurately reproducing the overall number of deaths. Additionally, accounting for the higher biological risk faced by males is essential for capturing gender-specific mortality trends. We also observe that because men had higher mortality rates, the models naturally prioritized fitting male data, sometimes at the expense of accuracy for females. Our work demonstrates that gender-agnostic models may miss crucial details. We conclude that incorporating gender-specific data is vital for creating more realistic models and designing inclusive public health policies that protect all demographic groups effectively.

## Introduction

Gender and sex are important, yet often overlooked, dimensions affecting the dynamics of infectious diseases [1]. From a socio-behavioral perspective, a consistent finding across studies is that females are more likely than males to adopt and maintain protective behaviors, including wearing face masks [2, 3], practicing social distancing [2, 3], and maintaining good hand hygiene [2–5]. Similar trends have been reported for COVID-19, particularly with regard to non-pharmaceutical interventions (NPIs), with females more likely than males to adopt NPIs and maintain them over time [6–8]. Broader societal gender roles also shape contact patterns through factors such as occupational segregation and caregiving responsibilities [9]. However, these patterns are not always straightforward; for example, females have shown significantly greater hesitancy to vaccine for several diseases, including COVID-19 [10–12]. From a biological perspective, research suggests that females often exhibit stronger immune responses than males [13, 14]. For example, the prevalence of hepatitis A and tuberculosis is higher in males than in females [15]. Similar patterns have been observed for COVID-19, with males showing a higher risk of developing severe illness and experiencing higher mortality rates [8, 16–19].

The recognized importance of sex and gender in shaping behaviors relevant to infectious disease transmission calls for their inclusion in epidemic models together with other variables that are already often taken into account. Age, in particular, is widely understood as being a key stratifying variable due to substantial differences between different age groups, not only regarding vulnerability to diseases but also in terms of contact patterns and behavioral responses [20–25]. Recently, research has also highlighted the need to integrate the diversity of social and structural conditions into models [26–28]. For instance, recent work has shown how generalized contact matrices, stratified by age and other variables such as socio-economic status (SES), allow developing expressive models able to correctly capture attack rates and disease burden across groups of the population [29]. This highlights how neglecting key social variables can lead to substantial misrepresentations of transmission dynamics, and motivates a shift towards models integrating a population stratified along multiple dimensions, and in particular towards the use of multi-dimensional contact matrices to feed data-driven models.

Although gender is among the most common socio-demographic variables collected in surveys investigating knowledge, attitudes, and behaviors toward infectious diseases [30, 31], sex and gender have rarely been explicitly incorporated into epidemic models [32, 33]. Analyses of large-scale contact surveys, such as POLYMOD [22] and CoMix [34, 35], have often been limited to exploring aggregate differences in the total number of contacts between males and females, without investigating deeper structural variations in their contact patterns. One of the few studies to construct and analyze generalized contact matrices that include gender is found in the COVID-19 literature [36]. However, the research used pre-pandemic data and its application to epidemic models required adjusting the matrices for the COVID-19 period in a manner that, while plausible, was not empirically validated. This example highlights a broader trend: while theoretical frameworks able to take into account gender differences have been proposed, practical applications remain limited [8, 37], probably due to the limited availability of gender-stratified data for epidemiological parameters and outcomes [38],.

Here, we address this gap by explicitly incorporating gender and gender-specific characteristics into age-stratified epidemic models in a data-informed manner. The goal is to identify which gender-related features have to be taken into account for capturing observed dynamics. In doing so, we consider features more closely associated with biological sex (understood as physiological and anatomical differences between males and females) alongside others more related to gender (understood as the socially and culturally constructed roles, behaviors, and norms associated with being male or female). Although these dimensions are conceptually distinct, we use the term “gender” throughout to refer to both for brevity and, due to data limitations, we restrict our representation to a binary classification. Building on a baseline age-structured Susceptible-Latent-Infectious-Recovered (SLIR) framework, extended to include deaths, we consider three gender-specific features that can be selectively introduced into the model: (1) a contact matrix stratified by both age and gender, (2) age- and gender-specific infection fatality ratios (IFRs), and (3) a behavioral component reflected in higher transmission rates among males compared to females. Each of these features can either be taken into account in the model or not. Eight different models can thus be built, each corresponding to a different way of accounting for differences between genders. These models range from a purely age-stratified model to one fully stratified by both age and gender for contacts as well as IFRs, and by gender for transmission rates. To assess the relative importance of these features, we consider these models in the context of the second wave of the COVID-19 pandemic in Italy (Autumn 2020 – Winter 2020*/*2021). More precisely, our methodology involves a two-step process. First, we calibrate all eight models against weekly death data aggregated by both age and gender. Second, we evaluate how well these calibrated models capture the empirically observed dynamics, not only on the aggregated data but also when the data is disaggregated by gender, by age, and by both age and gender.

Results show that including gender-specific IFR and contact patterns improves how well the models reproduce observed mortality data. In particular, we find that accounting for gender-specific IFR is key to properly capture the gender disaggregated mortality, and in particular the fact that male mortality is higher than female mortality at all times. Moreover, including gender-stratified contacts improves the performance of the model in reproducing not only empirical mortality disaggregated by both age and gender, but also mortality data disaggregated only by age. This suggests that taking into account gender differences in contact patterns can improve overall models’ realism. Our study also finds unexpected trade-offs: a model that captures well male mortality data in a certain age range may not reproduce as well female mortality data in another age range. This shows that a single “one-size-fits-all” model might not always be obtainable, and that models might need to be tailored to the specific groups and outcomes being studied.

## Materials and methods

### Data

In this study, we use data from multiple sources to stratify the population by age and gender, construct age- and gender-stratified contact matrices, and to calibrate the models.

**Contact matrices** We rely on two different sources to obtain contact matrices: the POLYMOD study [22] and the CoMix study [34, 39]. POLYMOD was a large-scale European survey conducted in 2006 that provides a pre-pandemic baseline for social mixing patterns, whereas CoMix was specifically designed to measure contact behaviors in several European countries during the COVID-19 pandemic. This dual-source approach is necessary because, on November 6, 2020, the Italian government implemented a tiered restriction system, assigning each of the 20 regions a daily color code—yellow, orange, or red—based on the local epidemiological situation. Each color corresponded to a different level of restrictions, with yellow indicating the lowest level and red the highest [40] (note that we do not consider the “white zone”, which technically corresponded to an even lower level of restrictions than the yellow zones, as white zones almost never occurred during the data collection period). The CoMix data collection waves occurred entirely during this period and are therefore not suitable to feed into models of the spread taking place before November 6, 2020, for which we thus use matrices derived from the POLYMOD study.

The first part of the procedure to obtain the age- and gender-stratified contact matrices from the data of the two studies is similar. We construct four separate age-stratified contact matrices: **c***^M,M^* for male-to-male contacts, **c***^M,F^* for male-to-female contacts, **c***^F,M^* for female-to-male contacts, and **c***^F,F^* for female-to-female contacts. The element (*i, j*) of each matrix represents the average number of contacts reported in the survey data between individuals of the corresponding gender(s) in age groups *i* and *j*. The matrices **c***^M,M^* and **c***^F,F^* are symmetric, i.e., 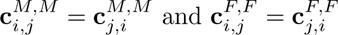. For matrices 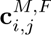 and 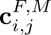 it holds 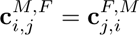, as the number of male-to-female contacts from age group *i* to age group *j* matches the number of female-to-male contacts from *j* to *i*. We then convert the total numbers of contacts to per capita numbers of contacts, by dividing each row of each matrix by the number of individuals in the corresponding age and gender group. Specifically, we divide row *i* of **c***^M,M^* and **c***^M,F^* by the number 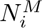 of males in age group *i*, and row *i* of **c***^F,F^* and **c***^F,M^* by the number 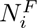 of females in age group *i*. The resulting matrices are not symmetric, as they now reflect the per capita contact rate rather than total contacts. Finally, we merge the four matrices into a single generalized contact rate matrix **c**, stratified by both age and gender, in which the entry **c***_ig,i_′_g_′* represents the mean contact rate for an individual in age group *i* and of gender *g* with individuals in age group *i^′^* and of gender *g^′^*.

Moreover, in a second step we take into account the impact of non-pharmaceutical interventions by modulating the contact matrices over time. This procedure depends on the specific dataset used (either POLYMOD or CoMix), as described in the following two paragraphs.

**POLYMOD data** When using the POLYMOD data, we compute a generalized contact matrix for five different settings: home, school, workplace, leisure, and general community setting. Variations in contact patterns induced by non-pharmaceutical interventions in the workplace, leisure, and general community setting are modeled using data from the Google COVID-19 Community Mobility Reports [41]. These reports provide the daily percentage *p_s_*(*t*) of visits to setting *s*, relative to a pre-pandemic baseline. A reduction factor which captures contact variation can thus be estimated as 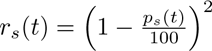, based on the assumption that the number of contacts is proportional to the square of the number of individuals present. Note that, while both Google Mobility data and POLYMOD data distinguish contacts by setting, their categorizations do not align perfectly. In some cases, a direct correspondence can be established, while in others an approximate mapping is required. The setting-specific parameters are derived as follows:

- For the POLYMOD *workplace* setting, we use the *workplaces percent change from baseline* field of Google Mobility data.
- For the POLYMOD *leisure* setting, we use the *retail and recreation percent change from baseline* field of Google Mobility data.
- For the POLYMOD *general community* setting, we use the average of the *grocery and pharmacy percent change from baseline* and the *transit stations percent change from baseline* fields of Google Mobility data.

To smooth out fluctuations in daily values, we compute the weekly average for each parameter.

In the case of schools, we rely on data from the Oxford COVID-19 Government Response Tracker [42], which provides a daily ordinal index *P* (*t*) (ranging from 0 for no measures to 3 for full closure) indicating the stringency of school closure policies at the national level. We compute the weekly average of this index and derive a school contact reduction factor as 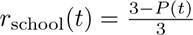. Finally, we assume that the home contact matrix remains constant, with no reduction factor applied. Indeed, while some studies have documented changes in household contacts in the case of strict lockdowns [43–45], the period during which we describe contacts with POLYMOD matrices did not include such measures in Italy.

The contact matrix at time *t* (where *t* is measured in weeks) is computed as a weighted combination of the five setting-specific matrices, each modulated by the corresponding reduction factor.

**CoMix data** We use the CoMix data to generate a separate contact matrix for each of the three restriction zones (i.e.,yellow, orange, and red) by considering only data from survey participants under the corresponding restriction zone. The contact matrix for a given day is then obtained as a weighted sum of the three zone-specific matrices, where the weights correspond to the proportion of the Italian population living under each restriction regime on that day:

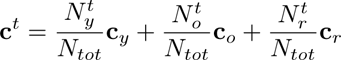

where **c***_y_*, **c***_o_*, **c***_r_* are the three contact matrices for, respectively, yellow, orange and red zones, 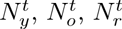 are the number of individuals in, respectively, yellow, orange, and red zones at time *t*, and *N_tot_* is the total population. Population data for each region, used to compute the weights, are taken from the Italian National Institute of Statistics (ISTAT) [46].

**Demography data** To stratify the population in our models, we use demographic data for Italy from the United Nations World Population Prospects [47]. We consider five age groups: 20–29 years old (y.o.), 30–39 y.o., 40–49 y.o., 50–59 y.o., and 60+ y.o., and two genders: male and female. Individuals under 20 years of age are excluded because of missing gender information for children (18 y.o. or less) and to maintain compatibility with the available mortality data used for model calibration, which are aggregated into 10-year age groups.

**Epidemiological data** For calibration, we use weekly death counts provided by the Italian National Institute of Health (ISS) [48] for the period corresponding to the second wave of COVID-19 in Italy. Specifically, the dataset covers the weeks from September 14, 2020 (Week 38) to February 21, 2021 (Week 7). These data are stratified by age (in 10-year intervals) and gender (male and female).

### Epidemic models

**Core structure of the model** We use a stochastic compartmental model with six compartments, illustrated in Fig 1. Susceptible individuals (*S*) become infected and move into the Latent compartment (*L*) at a rate known as the force of infection (*λ*), characterized by the transmission rate (*β*) and the contact matrix. In the latent stage, individuals are not yet infectious; they progress to the Infectious compartment (*I*) at rate *ɛ*. Infectious individuals leave the infectious state at rate *µ^−^*^1^: they either recover (*R*) or die (*D*), with respective probabilities given by the IFR, which is age-stratified in all model versions. To model more accurately the real-world delay between when a death occurs and when it is officially recorded, we introduce an intermediate compartment, *D*. Individuals that die from the infection first move from *I* to *D*, where they remain for an average duration Δ before being counted in the cumulative deaths compartment, *D°* (the superscript stands for “observed”). For the full set of equations that rule the model, see the Supplementary Material.

**Fig 1.**
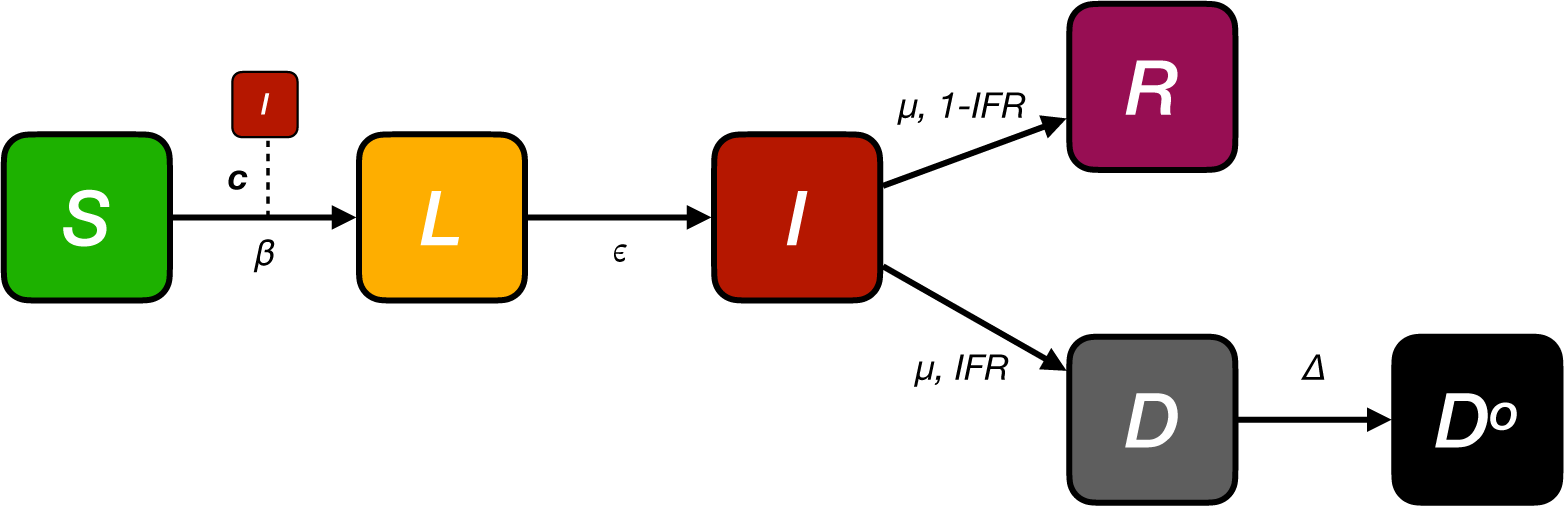
Schematic representation of the model. The model extends a standard SLIR (Susceptible–Latent–Infectious–Recovered) framework by including two compartments to account for deaths (*D* and *D°*). *D* represents an intermediate compartment to account for the delay between a death event and its official record. The contact matrix **c** used in the model are derived either from POLYMOD or CoMix, depending on the simulation period, and are stratified by age only or by both age and gender, depending on the model considered.

**Models with gender-stratified features** To investigate the impact of taking into account gender-stratified characteristics on epidemic outcomes, we define eight versions of the model, obtained from all possible combinations of three features: i) age- and gender-stratified contact matrix — denoted **c***_k,k_′* if stratified by age only, or **c***_kg,k_′_g_′* if stratified by both age (*k*) and gender (*g*); ii) age- and gender-stratified IFR — derived from the age-stratified IFR taken from the literature [49] and the odds ratio (OR = 1.39) reported in [50], representing the ratio of the odds of death between infected males and females. To find the specific Infection Fatality Rates for males (*IFR_m_*) and females (*IFR_f_*) for each age group, we solve the following system of equations:

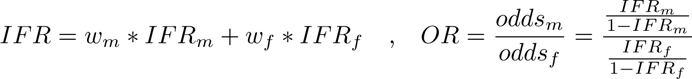

where *w_m_* and *w_f_* are the proportions of infected males and females, approximated by their proportions in the population for each age group; and iii) gender-specific behavior — represented by a multiplicative factor *r_β_ >* 1 applied to *β* for males, accounting for a higher risk of infection due to behavioral differences other than contacts.

To facilitate the analysis and the presentation of results, we assigned to each of the eight models a unique code based on the presence of the three gender-dependent features: contacts (C), IFR (I), and behaviors (B). The code 0 is used for the model that does not include any gender-dependent features. Models with multiple features combine the corresponding letters. The codes are reported in Table 1.

**Table 1.** Overview of model versions. Codes assigned to each of the 8 model versions based on the gender-dependent features included. Model 0 does not include any gender-dependency, while e.g. model CI uses generalized age- and gender-stratified contact matrices and an age- and gender-dependent IFR, but does not include the multiplicative factor *r_β_*.

| Model code | Contacts | IFR | Behaviors |
| --- | --- | --- | --- |
| 0 | ✗ | ✗ | ✗ |
| C | ✓ | ✗ | ✗ |
| I | ✗ | ✓ | ✗ |
| B | ✗ | ✗ | ✓ |
| CI | ✓ | ✓ | ✗ |
| CB | ✓ | ✗ | ✓ |
| IB | ✗ | ✓ | ✓ |
| CIB | ✓ | ✓ | ✓ |

**Parameters** The full list of parameters and their sources is reported in Table 2. The parameters *ɛ* [51, 52], *µ* [53, 54], and the age-stratified IFR [49] are taken from the literature, while for Δ we use a value of 14 days [55].The remaining parameters are fitted as described in the following section. Two values of *β* are estimated: *β*_1_ applies at the start of the simulation, and *β*_2_ applies from November 6, 2020, onwards, to account for stricter NPIs enforced on that date. Accordingly, POLYMOD contact matrices are used before November 6, 2020, and CoMix matrices thereafter, to account for the change in contact patterns.

**Table 2.** Summary of model parameters. Model parameters, their symbols, values, and sources from the literature.

| Parameter | Symbol | Value | Source |
| --- | --- | --- | --- |
| Transmission rate<br>(beginning of simulation) | $\beta_1$ | fitted | [58, 59] |
| Transmission rate<br>(6th November 2020) | $\beta_2$ | fitted | [58, 59] |
| Inverse of the latent period | $\epsilon$ | 0.25 days<sup>-1</sup> | [51, 52] |
| Inverse of recovery period | $\mu$ | 0.4 days<sup>-1</sup> | [53, 54] |
| Infection fatality ratio | IFR | stratified | [49, 50] |
| Delay in deaths | $\Delta$ | 14 days | [55] |
| Initial number of individuals distributed<br>in the infected compartments | $i_0$ | fitted | [60] |
| Initial fraction of<br>recovered individuals | $r_0$ | fitted | [60–62] |

The basic reproduction number *R*_0_ of all models is computed using the next generation matrix method [56, 57]. For more details on the formula of *R*_0_ for each model, see the Supplementary Material.

### Calibration

**Approximate Bayesian Computation** To calibrate our models, we use an Approximate Bayesian Computation (ABC) rejection algorithm [63, 64]. For additional details on the algorithm, see the Supplementary Material.

The free parameters of our models and their priors are:

- The two transmission rates, *β*_1_ and *β*_2_. For *β*_1_, we explore uniform values such that the corresponding *R*_0_ (*R*_01_) lies between 1 and 1.7. Although the basic reproduction number of COVID-19 was higher [58] early in the pandemic, several restrictions were already in place in Italy during Autumn 2020, justifying the use of smaller values. For *β*_2_, we explore uniform values such that the resulting *R*_0_ (*R*_02_) lies between 0.5 and 1.7. The reasoning is similar to that for *β*_1_, but we allow *β*_2_ to be below 1 due to the proximity to the epidemic peak. In both cases, the relevant metric is technically the effective reproduction number *R_t_*, given that there is a fraction of non susceptible individuals. However, given that the estimated initial recovered fraction *r*_0_ is relatively small (at most 10% of the total population) and the same goes for the fraction of infected individuals when we fit *β*_2_, we use the *R*_0_ as a suitable approximation for defining the prior distribution of the transmission rates *β*_1_ and *β*_2_.
- The initial number of infected individuals *i*_0_, assigned to the latent (L) and infectious (I) compartments based on the relative average time spent in each (*ɛ^−^*^1^ for *L* and *µ^−^*^1^ for *I*). We explore uniform values between the weekly number of confirmed cases reported in Italy on September 14, 2020 [60] (the start of the simulation – *i*_14*Sept*_ = 10, 119) and ten times that value.
- The initial fraction of recovered individuals *r*_0_. We explore values uniformly between 3% and 10% of the total population. The lower bound accounts for recoveries from the first COVID-19 wave, while the upper bound is based on two seroprevalence studies [61, 62] conducted in late 2020 in Italy, both reporting values around 10%. Since our simulation starts in September 2020, the fraction should be lower than the year-end estimates.
- The increased risk of infection for males *r_β_* due to behavioral differences, for the four models that include this feature. We consider a uniform prior distribution between 1 (no difference) and 1.3.

The models are calibrated against the weekly number of deaths aggregated by age and gender obtained from the ISS [48] between Week 40 of 2020 and Week 7 of 2021. Simulations start two weeks earlier (Week 38 of 2020) to allow the epidemic to evolve before calibration. For each model, we perform *N_sim_* = 10^6^ simulations and use as error metric *e* the weighted mean absolute percentage error (wMAPE) [65], defined as:

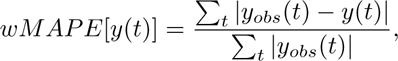

where *y_obs_*(*t*) and *y*(*t*) are respectively the observed and simulated weekly numbers of deaths for week *t*. We use a threshold value *σ* = 0.2 to determine acceptance or rejection of a parameter set.

**Evaluation metrics** For each model, we randomly draw 1, 000 parameter sets from the accepted posterior samples and run the model 10 times for each set, resulting in a total of 10,000 simulations per model. To assess how well each model explains the observed patterns, we use the root mean squared error (RMSE) to compare the mean predicted deaths across the 10,000 simulations against the observed death time series. To determine if the performance differences between models are statistically significant, we also compute an RMSE value for each individual simulation, yielding a distribution of 10,000 RMSE values per model. We then compare these distributions using a Kruskal-Wallis test followed by a Dunn’s post-hoc test. Furthermore, while the models are fitted using aggregated death data (total deaths across age and gender), we evaluate their ability to reproduce observed mortality at a more fine-grained level. To this end, we compute the RMSE on weekly death time series disaggregated by gender, by age, and by both gender and age. This approach allows us to quantify model performance in capturing mortality dynamics within specific demographic subgroups. Finally, to ensure the robustness of our findings, we evaluate model performances using an additional metric: the mean absolute error (MAE).

## Results

### Gender contacts in CoMix study

Although CoMix data has been studied in detail in other works [35, 66–69], gender-specific contact patterns have not been explored in depth. Previous analyses focused primarily on the total number of contacts of males and females, revealing only minor differences [35]. However, while the overall number of contacts is similar, the underlying interaction patterns can vary significantly between genders, and these differences can be visualized using contact matrices. To improve readability, instead of displaying the generalized contact matrix for each zone, in Fig 2A we show the four gender-specific matrices built using all available CoMix contact data for Italy: male-male, male-female, female-male, and female-female interactions. The female-female contact matrix reveals more diverse inter-age mixing. Specifically, females report a higher number of contacts outside their own age group (off-diagonal elements), especially with individuals aged 60 and above. In contrast, the male-male matrix is more strongly diagonal, indicating that male contacts are more concentrated within similar age groups. For cross-gender interactions, which are not symmetric (as they are per-capita contacts), we observe assortative mixing along the main diagonal as well as contacts with older individuals. However, females tend to have higher values in the elements immediately to the right of the diagonal, indicating a slight tendency to interact with males in the next older age group. For males, the reverse pattern is observed, with slightly larger values to the left of the diagonal. Finally, the female-to-male contact matrix shows higher per-capita contact rates than the male-to-female matrix.

**Fig 2.**
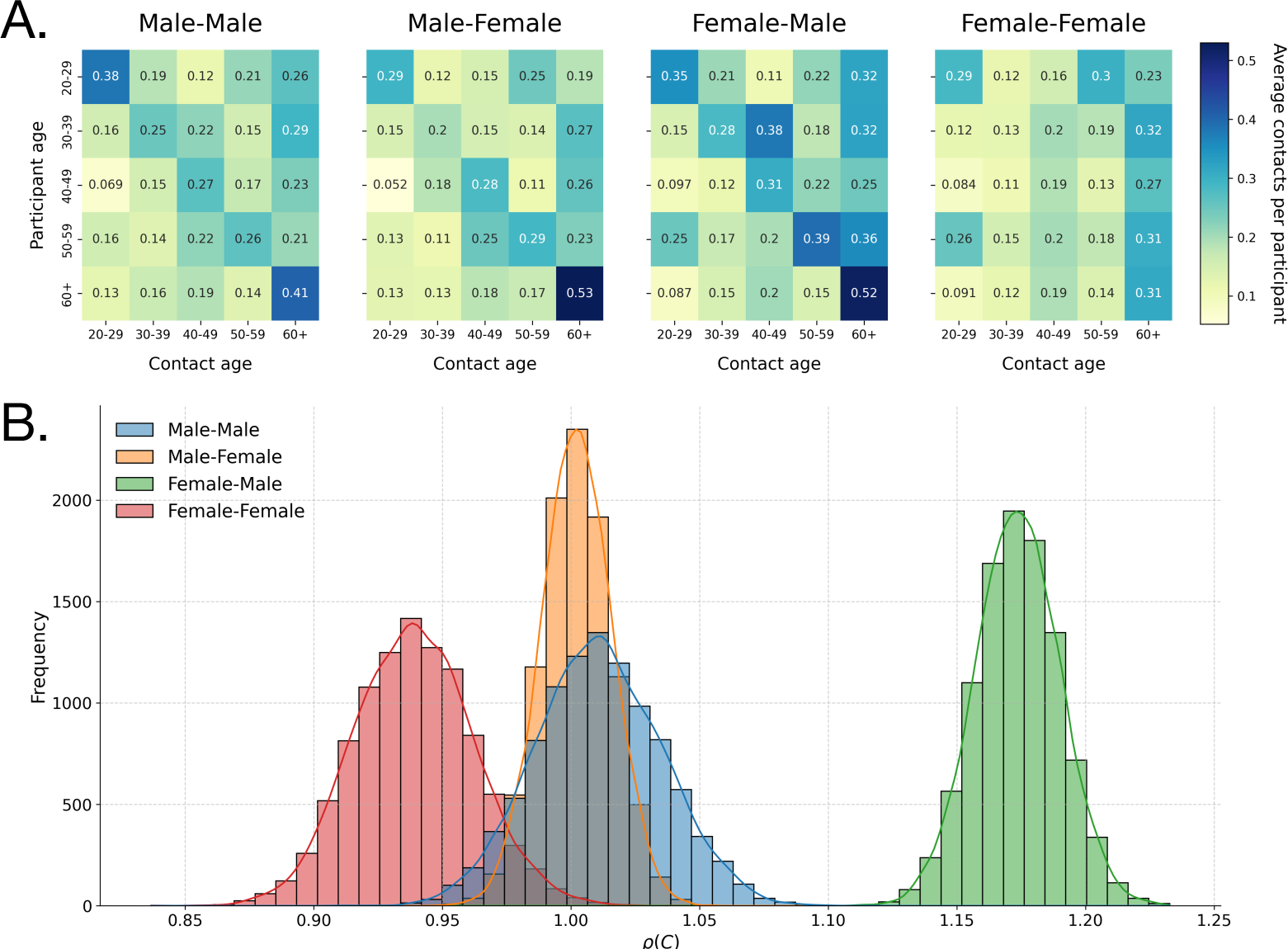
Gender-specific contact matrices and their dominant eigenvalues. A. Contact matrices for male-male, male-female, female-male, and female-female interactions, obtained from CoMix data for Italy. B. Distributions of the dominant eigenvalue of the four matrices, estimated via bootstrap with 10, 000 replicates for each matrix.

The epidemiological significance of these structural differences is captured by the dominant eigenvalue of each matrix, which is proportional to the potential for disease transmission (*R*_0_). This is illustrated in Fig 2B, which displays the distributions of the largest eigenvalue of each matrix, obtained via bootstrapping with 10, 000 replicates. We find that the female-female matrix has a lower dominant eigenvalue than the male-male one, suggesting that, in isolation, male-to-male contact patterns are more favorable to epidemic spread. Moreover, the female-to-male matrix has a substantially higher eigenvalue than its male-to-female counterpart. This implies a significant asymmetry in transmission potential: contact patterns make it more efficient for a disease to spread from females to males than from males to females. The Kolmogorov-Smirnov test applied to each pair of distributions confirms that the differences are statistically significant (*p <* 10*^−^*^4^). Given these findings, the remainder of our analysis is dedicated to investigating the impact of including this gender stratification, as well as the gender-dependency of IFR and behaviors, into epidemiological models.

### Models’ calibration

In Fig 3, we show the one-dimensional, marginal posterior distributions of *R*_01_, *R*_02_, *i*_0_, and *r*_0_ for each of the eight models, as well as the distribution of *r_β_* for the four models where it is included. We report the posterior distributions of *R*_01_ and *R*_02_ instead of *β*_1_ and *β*_2_ for ease of interpretation, given that the latter can be directly computed from the former.

**Fig 3.**
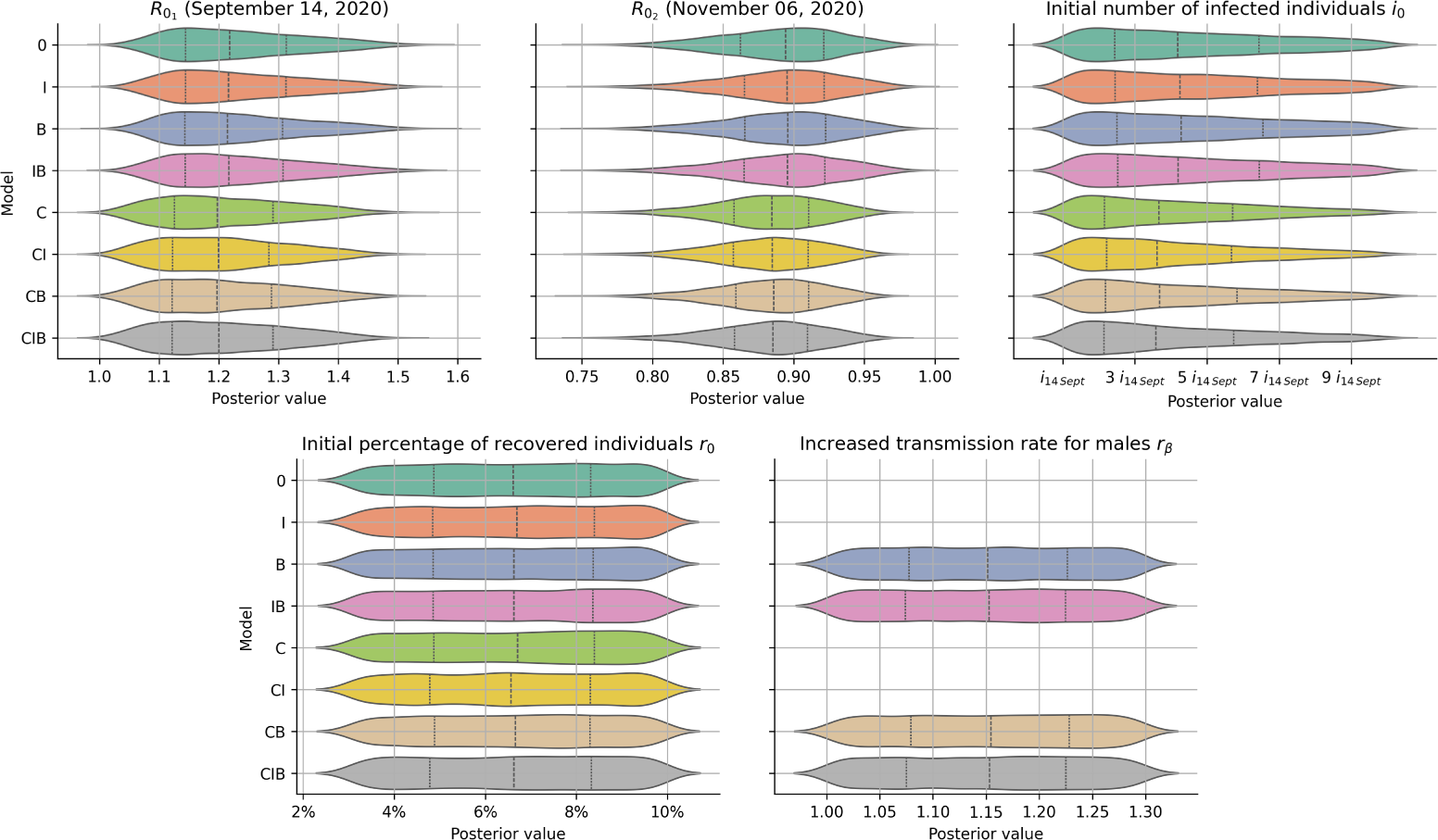
Posterior distributions of model parameters. Posterior distributions of *R*_01_, *R*_02_, *i*_0_, *r*_0_, and *r_β_* (when available) for each of the eight models (rows). The distributions are computed from all accepted parameter sets in the ABC rejection algorithm. Posterior values of *r*_0_ are expressed as percentages of recovered individuals out of the total population. Posterior values of *i*_0_ are expressed in units of *i*_14*Sept*_ which is the weekly reported number of infections on September 14, 2020, in Italy (10, 119). Model codes indicate which of the three features—contacts (C), IFR (I), and behaviors (B)—are present in each model (see Table 1).

Overall, the posterior distributions are quite similar across the different models. The mean values of *R*_01_ are above 1, as expected at the start of the simulations, while those of *R*_02_ are below 0.9, reflecting the implementation of control measures by November 6, 2020 and the proximity to the epidemic peak. Models with contacts stratified by gender tend to show posterior distributions for both *R*_01_ and *R*_02_ slightly shifted toward smaller values.

For *i*_0_, the posterior distributions tend to favor values close to the reported number of infections on September 14, 2020, although long tails are present. The posterior distributions of *r*_0_ and *r_β_* remain fairly uniform, similar to their priors. For *r*_0_, this suggests that the exact fraction of recovered individuals at the start of the simulation has limited influence on the model’s overall results. For *r_β_*, although males have a higher risk of infection in the model, no single value of this increased risk is clearly preferred.

In Fig 4 we present the results of the fit for the eight models, showing the mean weekly number of deaths across 10, 000 simulations (solid lines with shaded confidence intervals) alongside observed data (points). All models capture the general trend of the observed data, but the Root Mean Squared Error (RMSE) values, reported in each panel, reveal quantitative differences in performance. In particular, the results clearly split the models into two distinct groups based on the inclusion of gender-stratified contacts. The four models incorporating this feature (i.e., the C, CI, CB, CIB models) perform significantly better (lower RMSE) than the four models without it (i.e., the 0, I, B, IB models). This finding is statistically significant (Kruskal-Wallis, *p <* 10*^−^*^4^, see Supplementary Material).

**Fig 4.**
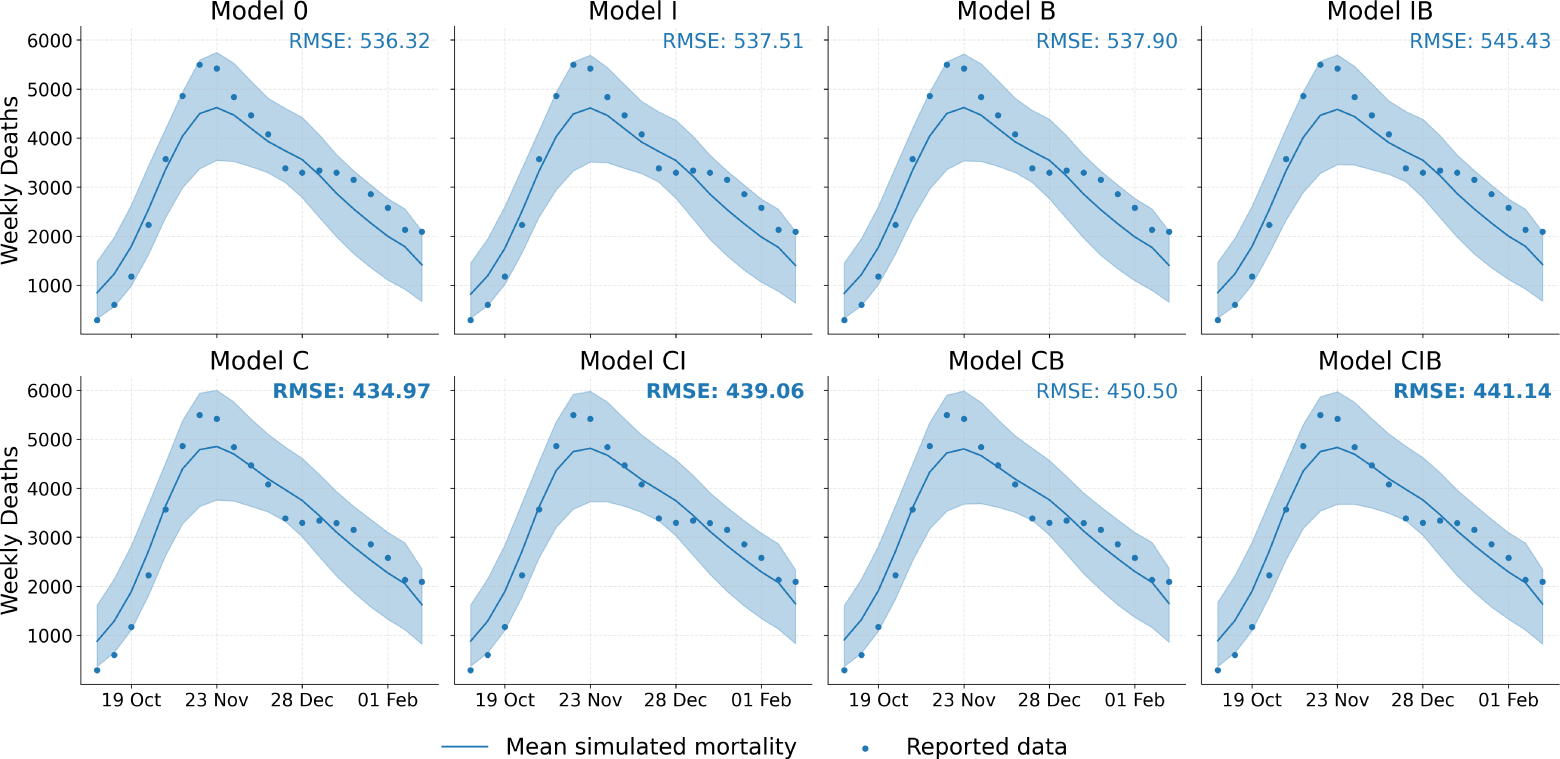
Model fit to aggregated mortality data. Simulated weekly number of deaths (mean and 95% confidence interval from 10,000 simulations; lines and shaded areas) compared with observed weekly numbers of deaths (dots) for each model. Root Mean Squared Error (RMSE) values are reported in each panel. The best (lowest) RMSE is highlighted in bold, as well as the two other RMSE values that are not statistically significantly different from the best (see Supplementary Material). Model codes indicate which of the three features—contacts (C), IFR (I), and behaviors (B)—are present in each model (see Table 1).

The model with only gender-stratified contacts (C) achieves the lowest RMSE (435.0), followed closely by model CI with gender-stratified contacts and IFR (RMSE = 439.1) and model CIB with all three gender-dependent features (RMSE = 441.1). The post-hoc analysis shows (Supplementary Material) that the differences among these top three models (C, CI, and CIB) are not statistically significant. Conversely, the four models lacking the gender-stratified contact feature (0, I, B, and IB) perform substantially worse, with RMSE values between 536.3 and 545.4. As with the top-performing group, the differences within this group of lower-performing models are not statistically significant. These results are robust with respect to choosing MAE as error metric (see the Supplementary Material).

### Models’ comparison with gender-disaggregated data

Fig 5 compares model performances in reproducing the empirical weekly number of gender-specific deaths. Each plot shows the mean across 10, 000 simulations (solid lines with shaded areas marking the confidence intervals) alongside observed data (points).

**Fig 5.**
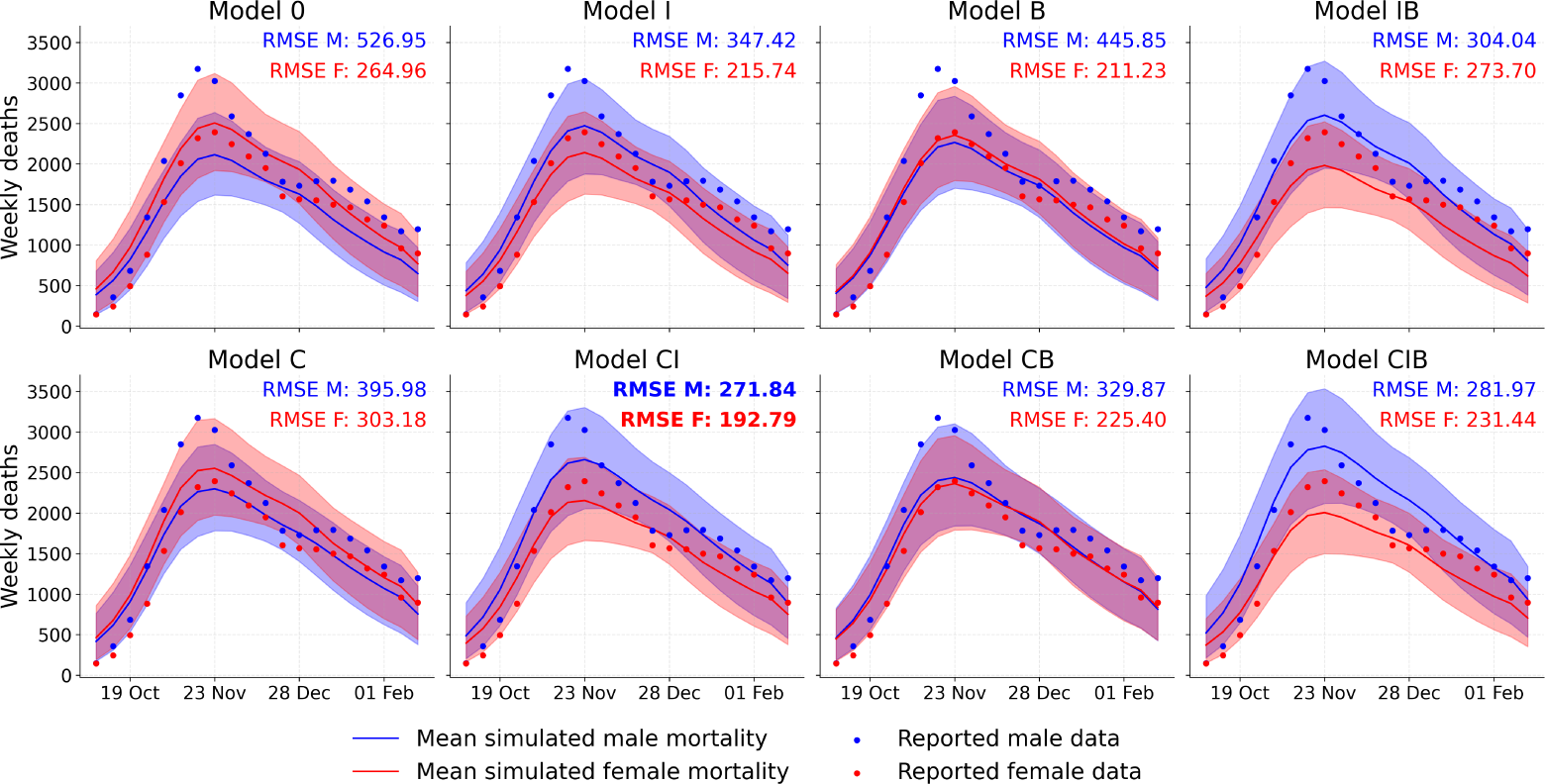
Comparison of model performance on gender-disaggregated mortality data. Weekly number of deaths obtained in the simulations (mean and 95% confidence interval from 10,000 simulations; lines and shaded areas), compared with the observed values (dots) for each of the eight models, stratified by gender. Parameter sets are drawn from the posterior distributions (1, 000 sets) and each set is simulated 10 times to account for stochastic variability. Root Mean Squared Error (RMSE) values for male (RMSE M) and female (RMSE F) mortality data are reported in each panel. The best (lowest) RMSE is highlighted in bold. Model codes indicate which of the three features—contacts (C), IFR (I), and behaviors (B)—are present in each model (see Table 1).

This analysis highlights the critical role of the gender-stratified IFR (models with I), particularly for capturing that the male mortality is larger than the female one. For male mortality data indeed, models lacking this feature (models without I, columns 1 and 3) tend to have high RMSE values, with the baseline model (0) having the highest (worst) RMSE (526.9). Moreover, male mortality is either lower than (models 0 and C) or similar to (models B and CB) female mortality, in qualitative disagreement with the empirical data. In contrast, models taking into account a gender-stratified IFR (models with I, columns 2 and 4) reproduce the qualitative trend of having a systematically larger male mortality. In addition, models in which gender-stratified IFR is combined to at least one other gender-related feature (models CI, IB, CIB) show a substantial improvement in RMSE values. The best RMSE is obtained for the model taking into account both gender-stratified contacts and gender-dependent IFR (model CI).

For female mortality data, the results are more complex. While the model incorporating both gender-stratified contacts and gender-dependent IFR (CI) also yields the best performance (lowest RMSE = 192.8), the other feature combinations show different effects. In particular, the model using only gender-stratified behaviors (B) achieves the second-best RMSE (211.2), performing slightly better than the model with only gender-stratified IFR (I) (RMSE = 215.7).

The tests presented in the Supplementary Material (Kruskal-Wallis, *p <* 10*^−^*^4^) show that all pairwise performance differences are statistically significant. Furthermore, also the analysis using the MAE as evaluation metric (see Supplementary Material) identifies model CI as the most robust and accurate model for reproducing gender-specific mortality data.

### Models’ comparison with age-disaggregated data

In Fig 6 we present the performance of each model in capturing the weekly number of deaths for each age group. All models overestimate the weekly number of deaths for the younger age groups (20 − 29 y.o. through 50 − 59 y.o.). However, the impact of this overestimation on the overall evaluation is limited, as the number of deaths in these groups is one to two orders of magnitude smaller than in the 60+ y.o. age group, which thus dominates the overall mortality signal.

**Fig 6.**
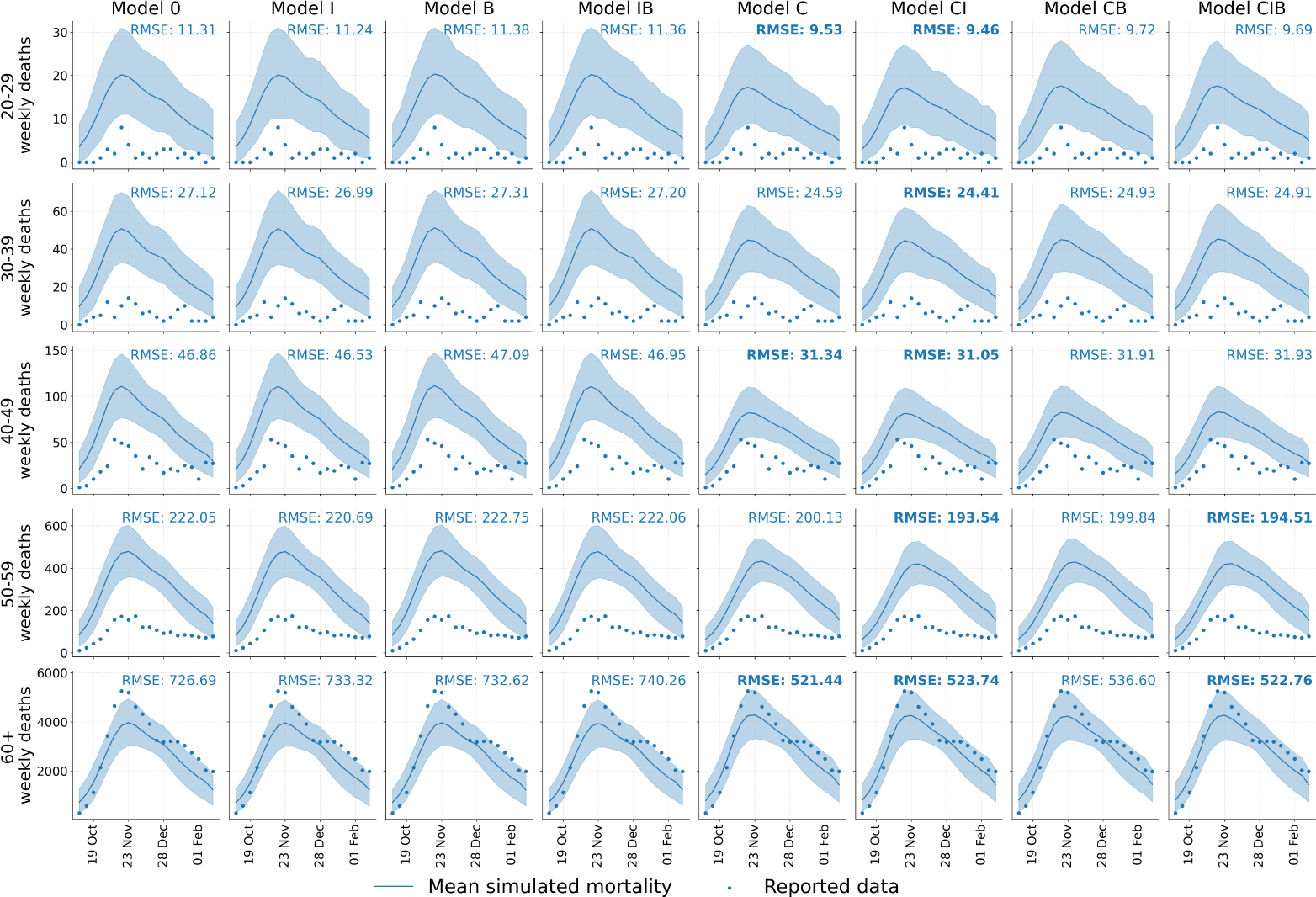
Comparison of model performance on age-disaggregated mortality data. Weekly number of deaths obtained in simulations (mean and 95% confidence interval from 10,000 simulations; lines and shaded areas) compared with observed values (dots) for each of the eight models (columns) and each age group (rows). Parameter sets are drawn from the posterior distributions (1, 000 sets) and each set is simulated 10 times to account for stochastic variability. Root Mean Squared Error (RMSE) values are reported in each panel. The best (lowest) RMSE for each age group is highlighted in bold, as well as the other RMSE values that are not statistically significantly different from the best (see Supplementary Material). Model codes indicate which of the three features—contacts (C), IFR (I), and behaviors (B)—are present in each model (see Table 1).

Similarly to the analysis on aggregated data, the comparison of the RMSE values obtained by the various models indicates that models using age- and gender-stratified contact matrices (models with C) reproduce age-disaggregated data systematically better than the other models. Indeed, these four C, CI, CB, CIB models have a lower (better) error than the four 0, I, B, IB models for each age group. This finding is statistically significant, as post-hoc tests confirm that every model with C performs significantly better than every model without C (*p <* 10*^−^*^3^ for all pairwise comparisons, see Supplementary Material). In fact, the four lower-performing models without C (0, I, B, IB) are largely indistinguishable from one another, particularly in the 40 − 49 y.o. and 50 − 59 y.o. age groups, for which no statistically significant differences are found between these models.

When comparing the top-performing models with C moreover, the model using both gender-stratified contact data and gender-dependent IFR (model CI) achieves the lowest RMSE for four of the five age groups (20 − 29 y.o., 30 − 39 y.o., 40 − 49 y.o., and 50 − 59 y.o.). For the 60+ y.o. age group, the model with gender-stratified contacts only (C) has the lowest RMSE (521.4), but its performance is statistically indistinguishable from the ones of model CIB (RMSE = 522.8) and of model CI (RMSE = 523.7). This suggests that for the older population, the addition of gender-stratified IFR or behavior features does not provide a significant improvement over the model taking only gender-stratified contact data into account. The analysis using the MAE as evaluation metric gives similar results (see Supplementary Material).

### Models’ comparison with age- and gender-disaggregated data

Finally, we analyze model performance at the most granular level, considering simultaneously both age and gender.In Fig 7, we show the curves of the weekly number of deaths for each model, age group, and gender, alongside the observed data and the corresponding RMSE values. This detailed view reveals a non-trivial pattern in which the best-performing model depends on the specific demographic group considered.

**Fig 7.**
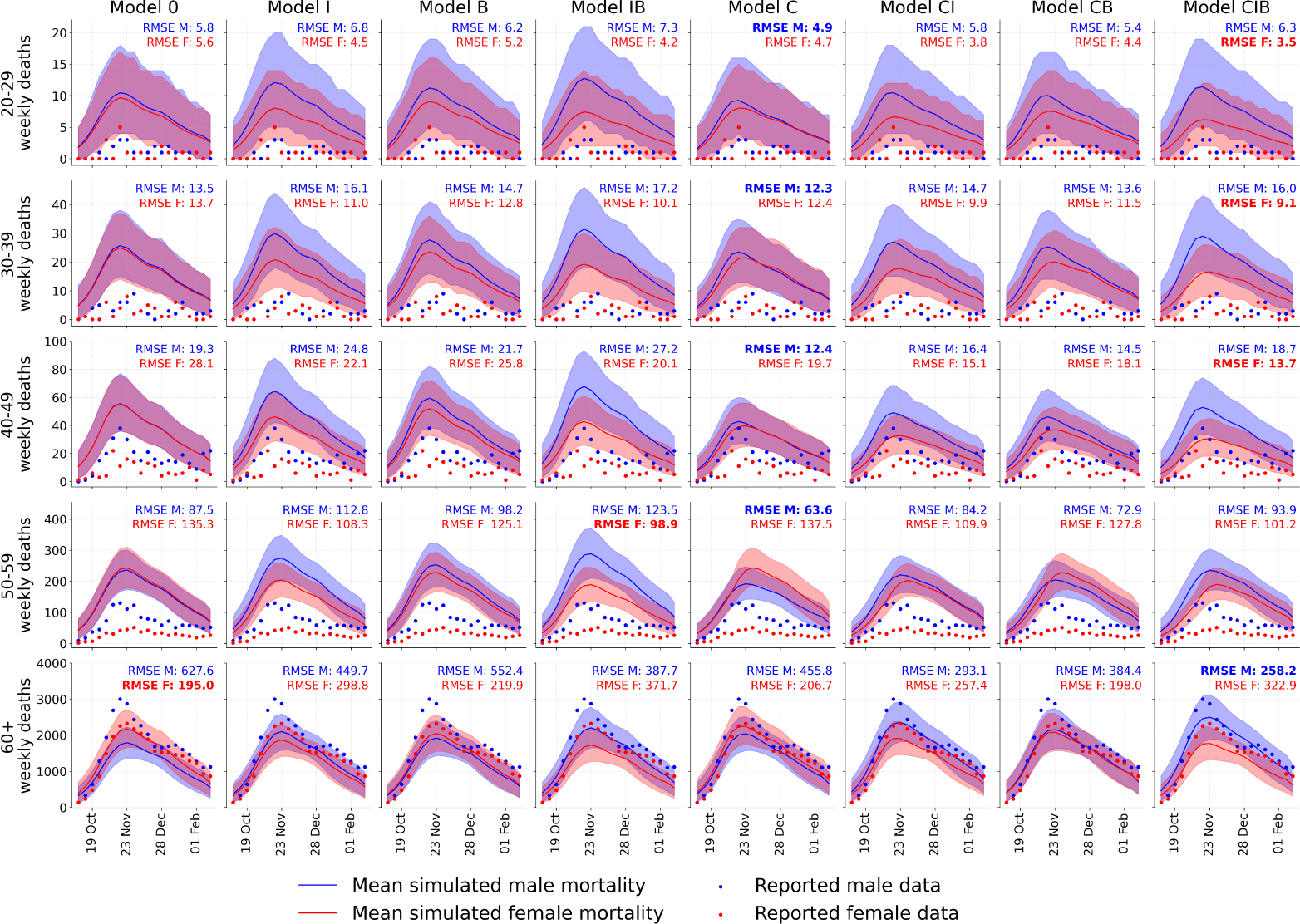
Comparison of model performance on age- and gender-disaggregated mortality data. Weekly number of deaths obtained in simulations (mean and 95% confidence interval from 10,000 simulations; lines and shaded areas) compared with observed values(dots) for each of the eight models (columns) and each age group (rows), stratified by gender. Parameter sets are drawn from the posterior distributions (1, 000 sets) and each set is simulated 10 times to account for stochastic variability. Root Mean Squared Error (RMSE) values for male (RMSE M) and female (RMSE F) mortality data are reported in each panel. The best (lowest) RMSE values for each age-gender group are highlighted in bold. Model codes indicate which of the three features—contacts (C), IFR (I), and behaviors (B)—are present in each model (see Table 1).

The results for the 60+ y.o. age group, which dominates the mortality signal, show differences between genders. For males over 60 y.o., the model incorporating all three gender-stratified features (CIB) performs best, achieving the lowest RMSE (258.19). It is followed by model CI with gender-stratified contacts and IFR (RMSE = 293.07). This indicates that accurately capturing mortality data in older males requires incorporating multiple gender-dependent features. On the contrary, for females over 60 y.o., the baseline model (0, with no gender-specific features) achieves the lowest RMSE (194.95). It is followed by models CB (RMSE = 198.01) and C (RMSE = 206.66). This finding suggests that for this demographic group, taking into account the standard age-specific IFR is sufficient to reproduce mortality data, and that adding the gendered features we considered actually worsens the performance.

The patterns in the younger age groups (20 − 59 y.o.) are also distinct. For males aged 20 − 59 y.o., the model with only gender-stratified contacts (C) is the best performer, achieving the lowest RMSE in all four age brackets. On the other hand, for females, the model with all gender-dependent features (CIB) is the top performer for the 20 − 29 y.o., 30 − 39 y.o., and 40 − 49 y.o. age groups. For the 50 − 59 y.o. age group, the model with gender-stratified IFR and behaviors (IB) provides the best performance.

Finally, and similarly to the analysis of age-aggregated mortality data, only models taking into account a gender-dependent IFR reproduce the fact that male mortality lies above female mortality across all age groups. While this trend is consistent in terms of total deaths, it is most visually evident in the groups above 40 y.o., as the significantly lower death counts in younger cohorts lead to greater fluctuations that can occasionally mask this pattern.

As shown in the Supplementary Material, the majority of these performance differences are statistically significant, underlining the clear demographic-specific nature of the epidemic spread. The analysis using MAE as performance metric confirms the robustness of our results (see Supplementary Material).

## Discussion

In this study, we investigated the effect of incorporating gender-related heterogeneities into an age-structured epidemic model of COVID-19 mortality in Italy. Despite the vast literature showing the existence of heterogeneities in disease-related behaviors [6, 7, 70–72] and in disease outcomes [16, 17], previous works have either focused on a theoretical characterization of frameworks [8, 37] or on contact matrices—adjusted in ways that are plausible but not empirically validated [36]. Our analysis of the gender-stratified contact matrices of the CoMix study revealed structural differences in contact patterns between genders, providing a strong incentive to investigate the impact of those patterns on the propagation of a disease in a population. Female-female contacts showed greater off-diagonal contacts, and female-to-male per-capita contacts were substantially higher than the reverse, a finding reinforced by the larger dominant eigenvalue of the female-to-male matrix. This observation of higher and more varied contacts among females is consistent with findings from the analysis by Doerre et al. [36] on the POLYMOD data for Germany. While their study used pre-pandemic data, our work confirms these structural patterns using data collected in Italy during the pandemic.

Starting from a SLIR framework including deaths, we thus built a series of data-driven model versions incorporating various combinations of age- and gender-stratified features. Namely, we considered a generalized contact matrix stratified by both gender and age, an age- and gender-dependent IFR and a phenomenological coefficient accounting for differences between genders in other disease-related behaviors. We first calibrated each model against weekly data collected during the second wave of the COVID-19 pandemic in Italy (September 14, 2020 to February 21, 2021), using data aggregated by both age and gender. We then conducted a comprehensive performance evaluation, first by assessing goodness of fit, and second by assessing their ability to reproduce mortality trends on disaggregated data (by gender only, by age group only, or by age and gender), that were not used to calibrate the model.

All eight models provided a qualitatively plausible fit of the aggregated mortality data. However, a quantitative comparison showed that models incorporating gender-stratified contact patterns (C, CI, CB, CIB) performed significantly better than those without. This initial finding suggests that contact patterns are the most dominant feature allowing to obtain a quantitative agreement with aggregated mortality data. Interestingly, the differences between the top-performing C-models (C, CI, and CIB) were not statistically significant, suggesting that incorporating gender-stratified IFR or behaviors into the model, in addition to gender- and age-dependent contact patterns, do not provide an additional benefit for the fit to aggregated mortality data.

This finding was reinforced by the analysis of the age-disaggregated data. Indeed, when mortality was disaggregated by age alone, models including gender-stratified contacts (C-models) outperformed the others across every age group. Among these, the model combining gender-stratified contacts and IFR (model CI) proved to be the most robust, achieving the best or statistically equivalent performance across all age categories. Hence, introducing gender-dependent contact patterns into a model can yield benefits that might extend beyond the description of gender-specific outcomes: indeed, incorporating this feature (contacts) improves the models’ ability to capture underlying age-based patterns.

Gender-stratified IFR also played a major role, and in particular taking them into account was necessary to reproduce the empirical higher mortality among males with respect to females. Moreover, this feature was most effective in improving performance when introduced in combination with gender-stratified contacts. Indeed, when disaggregating by gender alone, the model combining gender dependence in both contacts and IFR (CI) was the best performer for both males and females. This highlights the importance of including differences in mortality risk between genders, but also suggests that taking into account gender differences in biological vulnerability (IFR) alone is insufficient without also accounting for differences in contact patterns.

The role of the phenomenological description of differences in gender-stratified behaviors, such as adherence to NPIs or self-induced contact reduction, was more complex. Introduced in isolation (model B), it produced a good description of female mortality data (second-best model), while performing poorly for male mortality data. Moreover, in the study of mortality data fully disaggregated by age and gender, its inclusion was crucial for achieving the best performance in specific groups, namely to describe the mortality of older males (model CIB) and younger females (models CIB and IB).

Overall, the inclusion of gender-dependent features in the models tended to penalize the description of female mortality data in favor of reproducing male mortality. This trade-off is evident when looking at the different results for the 60+ y.o. age group in the fully disaggregated analysis. This group is by far the most impacted, with a substantially higher number of deaths. On the one hand, the baseline age-dependent model, with no gender-dependent features (0), captured best the mortality curve for females but substantially underestimated deaths among males, making it the worst-performing model in this regard. Moreover, the inclusion of all three gender-stratified features (model CIB) amplifies the shift of simulated data toward higher male mortality and lower female mortality, leading this model to achieve the best performance for male mortality data but the poorest performance for female mortality.

The sensitivity analysis done by using MAE as evaluation metric confirmed the robustness of our main conclusions, with the top-performing models in each analysis remaining consistent.

Our findings highlight that the impact of including gender-related heterogeneities in models is context-dependent: their contribution to model accuracy depends both on the epidemiological characteristics of the subgroup under consideration and on the nature of the heterogeneity being modeled. Even in contexts where outcome data are aggregated by gender—as was the case for the mortality data used to fit our models—incorporating gender-specific features can improve the realism of the simulations, particularly when the focus is on specific subpopulations. In practice, this suggests that models intended to inform public health interventions would benefit from incorporating gender-specific features, especially when aiming to evaluate impacts within subgroups where such differences are substantial.

The work presented in this study comes with several limitations. First and foremost, it is important to stress that our goal was not to develop a predictive model aimed at forecasting the pandemic trajectory. The model fitting was used to ground our simulations in data, but the primary aim was to understand the impact of gender differences in epidemic modeling, in a data-driven way, and to identify which features play the most prominent role in the results. Secondly, it is important to note that our approach uses the term “gender” to refer to a set of characteristics that, in reality, include both biological sex-related factors and socially constructed gender-related factors. This choice was made due to data limitations that preclude a complete separation of these concepts and to the lack of data allowing for a representation beyond the binary classification. Future work would benefit from data that enables a more nuanced treatment of sex and gender, including non-binary identities, as well as the disentangling of biological and social determinants of health. Thirdly, we could not include gender of children in our analysis, due to data availability. Despite the fact that the role of children in the transmission chain of the first wave of the COVID-19 in Italy is smaller than for other viruses such as influenza [73, 74], differences in caregiving patterns due to traditional gender roles may be a source of variation in exposure between males and females [9]. Thus, contact patterns including children would have been interesting to analyze and should be included in works of this kind. Additionally, in deriving the gender-specific IFRs, we weighted the contributions of males and females by their respective population shares rather than by the proportion of infections. While this assumption was necessary to ensure consistency with the aggregate age-stratified IFRs reported in the literature [49], it does not account for the fact that the ratio of infected males to females may dynamically deviate from the population ratio during the epidemic. Furthermore, we introduced behavioral differences not included in contact patterns in a deliberately simple way so as not to overcomplicate the framework or deal with an excessive number of fitting parameters. Future studies could explore more complex, ideally data-driven, representations of gendered behavioral differences. We also did not consider vaccination, as our simulations ended in February 2021 and COVID-19 vaccination in Italy began only in the very last days of December 2020, making its impact negligible in the first weeks of 2021. Nonetheless, incorporating vaccination in future models could be valuable, particularly in the light of documented gender differences in vaccine hesitancy [11, 12] and the impact of vaccination on contact behavior [39]. Additionally, a potential extension of this research would be to perform the model calibration directly on the disaggregated data. Our study was designed to fit models to aggregated mortality data and then evaluate their ability to explain disaggregated trends—a common scenario when detailed data is unavailable for calibration. By fitting directly to age- and gender-specific data, future analysis could determine whether the model selection outcomes change, revealing if the trade-offs observed here persist, and providing an even more granular understanding of which features are essential for specific demographic groups. Finally, our analysis focused exclusively on Italy. While this allowed for a detailed data-driven investigation, social contact patterns and gender roles vary across cultures. However, the framework developed here is general and can be readily applied to other countries where age- and gender-stratified data are available. Extending this analysis to other geographic contexts would be of great interest to investigate how the impact of gender-related variables on epidemic spread varies across different societies.

In conclusion, this study moves beyond the theoretical acknowledgment of gender disparities in infectious diseases to provide a systematic, data-driven evaluation of their impact within an epidemiological model. The results indicate that the inclusion of empirically grounded features—such as differences in contact patterns and in mortality risk—is not merely a refinement but can become a crucial step toward more accurate simulations. Our findings underscore that the path to improving epidemiological models lies in the careful, context-specific integration of key demographic and social structures.

## Data Availability

The aggregated mortality data used for calibration are publicly available from the Istituto Superiore di Sanita (ISS) (https://www.epicentro.iss.it/coronavirus/sars-cov-2-sorveglianza-dati). Google Community Mobility Reports are available from Google (https://www.google.com/covid19/mobility/). The CoMix contact data for Italy are available on Zenodo (https://doi.org/10.5281/zenodo.6362888). All other relevant data underlying the findings are within the manuscript and its Supporting Information files.

## Acknowledgments

ADG and DP acknowledge support from the Lagrange Project of the Institute for Scientific Interchange Foundation (ISI Foundation) funded by Fondazione Cassa di Risparmio di Torino (Fondazione CRT). AB acknowledges support from the Agence Nationale de la Recherche (ANR) project DATAREDUX (ANR-19-CE46-0008). The authors gratefully acknowledge Alberto Uriales and Antonino Bella from the ISS (Istituto Superiore di Sanita) for sharing the aggregated and disaggregated mortality data used in this study.

