## Supplementary Material for "Including gender-specific features in epidemic modeling: the case of the second wave of COVID-19 in Italy"

### S1 Model information

#### S1.1 Equations and simulations

The following set of differential equations describe the mean-field dynamics of model CIB for each of the six compartments (S,L,I,R,D,D0), specified for each gender $g$ and each age group $k$ :

$$\begin{aligned}
\frac{dS_{kg}}{dt} &= -r_{\beta_g}\beta \sum_{k'=1}^{K}\sum_{g'=1}^{G} \mathbf{c}_{kg,k'g'}\frac{I_{k'g'}}{N_{k'g'}}S_{kg} \\
\frac{dL_{kg}}{dt} &= r_{\beta_g}\beta \sum_{k'=1}^{K}\sum_{g'=1}^{G} \mathbf{c}_{kg,k'g'}\frac{I_{k'g'}}{N_{k'g'}}S_{kg} - \epsilon L_{kg} \\
\frac{dI_{kg}}{dt} &= \epsilon L_{kg} - \mu I_{kg} \\
\frac{dR_{kg}}{dt} &= \mu(1 - IFR_{kg})I_{kg} \\
\frac{dD_{kg}}{dt} &= \mu IFR_{kg}I_{kg} - \frac{1}{\Delta}D_{kg} \\
\frac{dD^o_{kg}}{dt} &= \frac{1}{\Delta}D_{kg}
\end{aligned} \tag{1}$$

Where $K$ is the total number of age groups and $G$ is the total number of gender groups in the population and $N_{k'g'}$ is the number of individuals of age k' and gender g'. The mean-field equations describing the other model versions are obtained by setting the corresponding terms to their gender-independent values (e.g., setting $r_{\beta_g} = 1$ for models without gender-specific behavior, or using a contact matrix $\mathbf{c}_{k,k'}$ that is only stratified by age).

To preserve the discrete and stochastic nature of transitions between compartments, we do not directly integrate these equations. Instead, we implement a discrete-time stochastic simulation with a time-step of one day. At each time $t$, the number of individuals in age group $k$, gender group $g$, that move from compartment $X$ to compartment $Y$ is sampled from:

$$Pr_{bin}(X_{k,g}(t), p_{X_{k,g}\to Y_{k,g}}(t)),$$

where $Pr_{bin}(N,p)$ is the binomial probability density function with $N$ trials and probability of success $p$ and $p_{X_{k,g}\to Y_{k,g}}(t)$ is the corresponding transition probability. This probability is derived from the rates in the continuous model (Eq. 1) for a time step of $\Delta t = 1$ day. Specifically, for a transition with a per-capita rate $\lambda$, the probability is calculated as $p = 1 - e^{-\lambda\Delta t}$. For example, the probability for a latent individual to become infectious is $p_{L\to I} = 1 - e^{-\epsilon\Delta t}$, while the probability for a susceptible individual in group $(k,g)$ to become infected is $p_{S\to L} = 1 - e^{-\lambda_{k,g}(t)\Delta t}$, where $\lambda_{k,g}(t)$ is the force of infection:

$$\lambda_{k,g}(t) = r_{\beta_g}\beta \sum_{k'=1}^{K}\sum_{g'=1}^{G} \mathbf{c}_{kg,k'g'}\frac{I_{k'g'}}{N_{k'g'}} \ .$$

#### S1.2 Basic reproduction number $R_0$

For all models without gender-stratified behavior, the basic reproduction number takes the form $R_0 = \frac{\beta}{\mu}\rho(\mathbf{T})$, where $\rho(\mathbf{T})$ is the spectral radius of the contact-related mixing matrix $\mathbf{T}$. Based on the model version, $\mathbf{T}$ is defined as $T_{kk'} = c_{kk'}\frac{N_k}{N'_k}$ if the contact matrix $\mathbf{c}$ is stratified only by age, or as $T_{kgk'g'} = c_{kgk'g'} \frac{N_{kg}}{N_{k'g'}}$ if the contact matrix $\mathbf{c}$ is stratified by both age and gender. For models incorporating gender-stratified behavior, where males have a transmission rate increased by a factor $r_\beta > 1$, the reproduction number is instead given by $R_0 = \frac{\beta}{\mu}\rho\left(\mathbf{D}_{r_\beta}\mathbf{T}\right)$, where $D_{r_\beta}$ is a diagonal matrix of dimension matching the one of $\mathbf{T}$, with diagonal entries equal to $r_\beta$ for male groups and 1 for female groups.

##### S1.3 ABC rejection algorithm

The ABC rejection algorithm we used to calibrate the models works as follows. We choose a prior distribution $P(\mathbf{\Theta})$ for the free parameters $\mathbf{\Theta}$, define an error metric $e$ to compare empirical and simulated time series of weekly deaths, and specify a total number of simulations $N_{sim}$ to perform. The algorithm samples a set of parameter values and runs a simulation to obtain a simulated time series $M'$ of weekly deaths. $M'$ is then compared to the empirical time series of weekly deaths $M$ using the error metric: if $e(M, M')$ is below the threshold $\sigma$, the parameter set is retained; otherwise, it is discarded. A new parameter set is sampled, and the process is repeated until the total number of simulations, $N_{sim}$, has been completed, regardless of acceptance or rejection. The collection of all retained parameter sets forms their joint posterior distribution.

#### S2 Statistical tests of model performances

As mentioned in the main article, to verify that the RMSE values computed for each model were statistically different from each other, we performed a Kruskal-Wallis test. In case of a p-value smaller than 0.05 and, thus significant differences among the RMSE values of the models, we followed up with a Dunn's post-hoc test using a Bonferroni correction, in which the statistical differences of each pair of models is checked. The results for the four different performances we analyzed is reported below. In all cases, the Kruskal-Wallis test was significant and, thus, we present only the results of the Dunn's post-hoc tests.

##### S2.1 Models' performances with aggregated data

The results of the Dunn's post-hoc test (Table S1) reveal a clear division of the models into two distinct groups: those without gender-stratified contacts (models 0, I, B, IB) and those with them (models C, CI, CB, CIB).

The performance difference between any model in the first group and any model in the second group is highly statistically significant ($p \leq 10^{-4}$ for all comparisons). Conversely, within each of these two main groups, the differences in RMSE are largely statistically insignificant ($p > 0.05$). This indicates that for modeling aggregated data, the inclusion of gender-stratified contacts is the dominant factor. The effects of gender-stratified IFR (indicated by I) and transmission rates (indicated by B) appear to be negligible.

##### S2.2 Models' performances with gender-disaggregated data

The results of the tests done to analyze the performances by gender are shown in Table S2. We can observe that for both males (top) and females (bottom), every model produces a statistically unique RMSE, with the vast majority of pairwise comparisons being highly significant ($p \leq 10^{-4}$). Therefore, all three gender-stratified features play a significant role in modeling gender-specific outcomes.

| | **0** | **I** | **B** | **IB** | **C** | **CI** | **CB** | **CIB** |
|---|---|---|---|---|---|---|---|---|
| **0** | 1.00 | 0.07 | 1.00 | 1.00 | **** | **** | **** | **** |
| **I** | 0.07 | 1.00 | * | **** | **** | **** | **** | **** |
| **B** | 1.00 | * | 1.00 | 1.00 | **** | **** | **** | **** |
| **IB** | 1.00 | **** | 1.00 | 1.00 | **** | **** | **** | **** |
| **C** | **** | **** | **** | **** | 1.00 | 1.00 | ** | 1.00 |
| **CI** | **** | **** | **** | **** | 1.00 | 1.00 | **** | 0.20 |
| **CB** | **** | **** | **** | **** | ** | **** | 1.00 | 1.00 |
| **CIB** | **** | **** | **** | **** | 1.00 | 0.20 | 1.00 | 1.00 |

Table S1: Results of the Dunn's post-hoc test for pairwise comparisons between models on the RMSE metric for the aggregated data. P-values are adjusted using the Bonferroni correction. Significance levels are indicated by: $****$: $p \leq 10^{-4}$; $***$: $10^{-4} < p \leq 10^{-3}$; $**$: $10^{-3} < p \leq 10^{-2}$; $*$: $10^{-2} < p \leq 0.05$. P-values greater than 0.05 are shown rounded to two decimal places.

##### S2.3 Models' performances with age-disaggregated data

The results for age-disaggregated data are shown in Table S3.

Similarly to the results for the aggregated data, the primary division based on gender-stratified contacts remains the single most dominant factor. Indeed, across all five age groups, every comparison between a model without contacts (models 0, I, B, IB) and a model with contacts (models C, CI, CB, CIB) is highly statistically significant ($p \leq 10^{-4}$). This reinforces that this feature is fundamental to model performance, regardless of age.

On the other hand, the analysis reveals other complex, age-dependent interactions. Within the group without contacts (0, I, B, IB), the models are largely statistically similar, especially for ages 40 and older. For younger ages (20-39), the gender-stratified behaviors show some significant impact (e.g., B vs 0 for 30-39), but this effect disappears in older age brackets. Within the group with contacts (C, CI, CB, CIB) a clear, age-dependent pattern emerges. For the 20-29, 30-39, and 40-49 age groups, the tables split into two new clusters based on gender-stratified behaviors. The performances of models C and CI (no behaviors) are statistically equivalent, and the same goes for models CB and CIB (with behaviors), but the two clusters are significantly different from one another. On the contrary, for the 50-59 age group, the split is based on the gender-stratified IFR. Models C and CB (no IFR) become statistically equivalent, as do CI and CIB (with IFR). Finally, for the 60+ group, models C, CI, and CIB are statistically similar, while CB (gender-stratified contacts and behaviors) stands out as significantly different from the others. In summary, while gender-stratified contacts are universally important, the relative importance of gender-stratified IFR and gender-stratified behaviors depends on the specific age group. The behavior feature appears more impactful for younger populations, while the IFR feature becomes more important for the 50-59 age group.

##### S2.4 Models' performances with age- and gender-disaggregated data

In Table S4, we show the results of the Dunn's post-hoc test for the metrics obtained comparing data disaggregated by both age and gender. For most age-gender combinations, nearly all models are highly statistically different from one another ($p \leq 10^{-4}$). This is particularly true for females in the 20-29, 30-39, and 40-49 age groups, and for males in the 40-49 and 50-59 groups. This means that at this granular level, every feature choice (contacts, IFR, behaviors) creates a model with a

**Males**

| | **0** | **I** | **B** | **IB** | **C** | **CI** | **CB** | **CIB** |
|---|---|---|---|---|---|---|---|---|
| **0** | 1.00 | **** | **** | **** | **** | **** | **** | **** |
| **I** | **** | 1.00 | **** | **** | **** | **** | **** | **** |
| **B** | **** | **** | 1.00 | **** | **** | **** | **** | **** |
| **IB** | **** | **** | **** | 1.00 | **** | **** | ** | * |
| **C** | **** | **** | **** | **** | 1.00 | **** | **** | **** |
| **CI** | **** | **** | **** | **** | **** | 1.00 | **** | **** |
| **CB** | **** | **** | **** | ** | **** | **** | 1.00 | **** |
| **CIB** | **** | **** | **** | * | **** | **** | **** | 1.00 |

**Females**

| | **0** | **I** | **B** | **IB** | **C** | **CI** | **CB** | **CIB** |
|---|---|---|---|---|---|---|---|---|
| **0** | 1.00 | **** | **** | **** | **** | **** | **** | **** |
| **I** | **** | 1.00 | **** | **** | **** | **** | **** | **** |
| **B** | **** | **** | 1.00 | **** | **** | **** | * | **** |
| **IB** | **** | **** | **** | 1.00 | **** | **** | **** | **** |
| **C** | **** | **** | **** | **** | 1.00 | **** | **** | **** |
| **CI** | **** | **** | **** | **** | **** | 1.00 | **** | **** |
| **CB** | **** | **** | * | **** | **** | **** | 1.00 | **** |
| **CIB** | **** | **** | **** | **** | **** | **** | **** | 1.00 |

Table S2: Results of the Dunn's post-hoc test for pairwise comparisons between models on the RMSE metric for the disaggregated data by gender (males on top and females on the bottom). P-values are adjusted using the Bonferroni correction. Significance levels are indicated by: ****: $p \leq 10^{-4}$; ***: $10^{-4} < p \leq 10^{-3}$; **: $10^{-3} < p \leq 10^{-2}$; *: $10^{-2} < p \leq 0.05$. P-values greater than 0.05 are shown rounded to two decimal places.

unique and statistically distinct performance.

**Age group 20-29**

| | 0 | I | B | IB | C | CI | CB | CIB |
|---|---|---|---|---|---|---|---|---|
| **0** | 1.00 | 1.00 | 0.09 | 1.00 | **** | **** | **** | **** |
| **I** | 1.00 | 1.00 | **** | * | **** | **** | **** | **** |
| **B** | 0.09 | **** | 1.00 | 1.00 | **** | **** | **** | **** |
| **IB** | 1.00 | * | 1.00 | 1.00 | **** | **** | **** | **** |
| **C** | **** | **** | **** | **** | 1.00 | 0.07 | **** | **** |
| **CI** | **** | **** | **** | **** | 0.07 | 1.00 | **** | **** |
| **CB** | **** | **** | **** | **** | **** | **** | 1.00 | 1.00 |
| **CIB** | **** | **** | **** | **** | **** | **** | 1.00 | 1.00 |

**Age group 30-39**

| | 0 | I | B | IB | C | CI | CB | CIB |
|---|---|---|---|---|---|---|---|---|
| **0** | 1.00 | 1.00 | ** | 1.00 | **** | **** | **** | **** |
| **I** | 1.00 | 1.00 | **** | * | **** | **** | **** | **** |
| **B** | ** | **** | 1.00 | 0.52 | **** | **** | **** | **** |
| **IB** | 1.00 | * | 0.52 | 1.00 | **** | **** | **** | **** |
| **C** | **** | **** | **** | **** | 1.00 | ** | **** | **** |
| **CI** | **** | **** | **** | **** | ** | 1.00 | **** | **** |
| **CB** | **** | **** | **** | **** | **** | **** | 1.00 | 1.00 |
| **CIB** | **** | **** | **** | **** | **** | **** | 1.00 | 1.00 |

**Age group 40-49**

| | 0 | I | B | IB | C | CI | CB | CIB |
|---|---|---|---|---|---|---|---|---|
| **0** | 1.00 | 1.00 | 1.00 | 1.00 | **** | **** | **** | **** |
| **I** | 1.00 | 1.00 | 0.05 | 1.00 | **** | **** | **** | **** |
| **B** | 1.00 | 0.05 | 1.00 | 1.00 | **** | **** | **** | **** |
| **IB** | 1.00 | 1.00 | 1.00 | 1.00 | **** | **** | **** | **** |
| **C** | **** | **** | **** | **** | 1.00 | 0.76 | *** | **** |
| **CI** | **** | **** | **** | **** | 0.76 | 1.00 | **** | **** |
| **CB** | **** | **** | **** | **** | *** | **** | 1.00 | 1.00 |
| **CIB** | **** | **** | **** | **** | **** | **** | 1.00 | 1.00 |

**Age group 50-59**

| | 0 | I | B | IB | C | CI | CB | CIB |
|---|---|---|---|---|---|---|---|---|
| **0** | 1.00 | 1.00 | 0.97 | 1.00 | **** | **** | **** | **** |
| **I** | 1.00 | 1.00 | ** | 1.00 | **** | **** | **** | **** |
| **B** | 0.97 | ** | 1.00 | 1.00 | **** | **** | **** | **** |
| **IB** | 1.00 | 1.00 | 1.00 | 1.00 | **** | **** | **** | **** |
| **C** | **** | **** | **** | **** | 1.00 | **** | 1.00 | **** |
| **CI** | **** | **** | **** | **** | **** | 1.00 | **** | 0.16 |
| **CB** | **** | **** | **** | **** | 1.00 | **** | 1.00 | **** |
| **CIB** | **** | **** | **** | **** | **** | 0.16 | **** | 1.00 |

**Age group 60+**

| | 0 | I | B | IB | C | CI | CB | CIB |
|---|---|---|---|---|---|---|---|---|
| **0** | 1.00 | 1.00 | 0.26 | * | **** | **** | **** | **** |
| **I** | 1.00 | 1.00 | 1.00 | 1.00 | **** | **** | **** | **** |
| **B** | 0.26 | 1.00 | 1.00 | 1.00 | **** | **** | **** | **** |
| **IB** | * | 1.00 | 1.00 | 1.00 | **** | **** | **** | **** |
| **C** | **** | **** | **** | **** | 1.00 | 1.00 | * | 1.00 |
| **CI** | **** | **** | **** | **** | 1.00 | 1.00 | ** | 1.00 |
| **CB** | **** | **** | **** | **** | * | ** | 1.00 | * |
| **CIB** | **** | **** | **** | **** | 1.00 | 1.00 | * | 1.00 |

Table S3: Results of the Dunn's post-hoc test for pairwise comparisons between models on the RMSE metric for the data disaggregated by age. P-values are adjusted using the Bonferroni correction. Significance levels are indicated by: ****: $p \leq 10^{-4}$; ***: $10^{-4} < p \leq 10^{-3}$; **: $10^{-3} < p \leq 10^{-2}$; *: $10^{-2} < p \leq 0.05$. P-values greater than 0.05 are shown rounded to two decimal places.

**Age 20-29 (Males)**

| | 0 | I | B | IB | C | CI | CB | CIB |
|---|---|---|---|---|---|---|---|---|
| **0** | 1.00 | **** | **** | **** | **** | 1.00 | **** | **** |
| **I** | **** | 1.00 | **** | **** | **** | **** | **** | **** |
| **B** | **** | **** | 1.00 | **** | **** | **** | **** | ** |
| **IB** | **** | **** | **** | 1.00 | **** | **** | **** | **** |
| **C** | **** | **** | **** | **** | 1.00 | **** | **** | **** |
| **CI** | 1.00 | **** | **** | **** | **** | 1.00 | **** | **** |
| **CB** | **** | **** | **** | **** | **** | **** | 1.00 | **** |
| **CIB** | **** | **** | ** | **** | **** | **** | **** | 1.00 |

**Age 20-29 (Females)**

| | 0 | I | B | IB | C | CI | CB | CIB |
|---|---|---|---|---|---|---|---|---|
| **0** | 1.00 | **** | **** | **** | **** | **** | **** | **** |
| **I** | **** | 1.00 | **** | **** | **** | **** | **** | **** |
| **B** | **** | **** | 1.00 | **** | **** | **** | **** | **** |
| **IB** | **** | **** | **** | 1.00 | **** | **** | **** | **** |
| **C** | **** | **** | **** | **** | 1.00 | **** | **** | **** |
| **CI** | **** | **** | **** | **** | **** | 1.00 | **** | **** |
| **CB** | **** | **** | **** | **** | **** | **** | 1.00 | **** |
| **CIB** | **** | **** | **** | **** | **** | **** | **** | 1.00 |

**Age 30-39 (Males)**

| | 0 | I | B | IB | C | CI | CB | CIB |
|---|---|---|---|---|---|---|---|---|
| **0** | 1.00 | **** | **** | **** | **** | **** | 0.76 | **** |
| **I** | **** | 1.00 | **** | **** | **** | **** | **** | **** |
| **B** | **** | **** | 1.00 | **** | **** | 1.00 | **** | **** |
| **IB** | **** | **** | **** | 1.00 | **** | **** | **** | **** |
| **C** | **** | **** | **** | **** | 1.00 | **** | **** | **** |
| **CI** | **** | **** | 1.00 | **** | **** | 1.00 | **** | **** |
| **CB** | 0.76 | **** | **** | **** | **** | **** | 1.00 | **** |
| **CIB** | **** | **** | **** | **** | **** | **** | **** | 1.00 |

**Age 30-39 (Females)**

| | 0 | I | B | IB | C | CI | CB | CIB |
|---|---|---|---|---|---|---|---|---|
| **0** | 1.00 | **** | **** | **** | **** | **** | **** | **** |
| **I** | **** | 1.00 | **** | **** | **** | **** | **** | **** |
| **B** | **** | **** | 1.00 | **** | **** | **** | **** | **** |
| **IB** | **** | **** | **** | 1.00 | **** | **** | **** | **** |
| **C** | **** | **** | **** | **** | 1.00 | **** | **** | **** |
| **CI** | **** | **** | **** | **** | **** | 1.00 | **** | **** |
| **CB** | **** | **** | **** | **** | **** | **** | 1.00 | **** |
| **CIB** | **** | **** | **** | **** | **** | **** | **** | 1.00 |

**Age 40-49 (Males)**

| | 0 | I | B | IB | C | CI | CB | CIB |
|---|---|---|---|---|---|---|---|---|
| **0** | 1.00 | **** | **** | **** | **** | **** | **** | **** |
| **I** | **** | 1.00 | **** | **** | **** | **** | **** | **** |
| **B** | **** | **** | 1.00 | **** | **** | **** | **** | **** |
| **IB** | **** | **** | **** | 1.00 | **** | **** | **** | **** |
| **C** | **** | **** | **** | **** | 1.00 | **** | **** | **** |
| **CI** | **** | **** | **** | **** | **** | 1.00 | **** | **** |
| **CB** | **** | **** | **** | **** | **** | **** | 1.00 | **** |
| **CIB** | **** | **** | **** | **** | **** | **** | **** | 1.00 |

**Age 40-49 (Females)**

| | 0 | I | B | IB | C | CI | CB | CIB |
|---|---|---|---|---|---|---|---|---|
| **0** | 1.00 | **** | **** | **** | **** | **** | **** | **** |
| **I** | **** | 1.00 | **** | **** | **** | **** | **** | **** |
| **B** | **** | **** | 1.00 | **** | **** | **** | **** | **** |
| **IB** | **** | **** | **** | 1.00 | **** | **** | **** | **** |
| **C** | **** | **** | **** | **** | 1.00 | **** | **** | **** |
| **CI** | **** | **** | **** | **** | **** | 1.00 | **** | **** |
| **CB** | **** | **** | **** | **** | **** | **** | 1.00 | **** |
| **CIB** | **** | **** | **** | **** | **** | **** | **** | 1.00 |

**Age 50-59 (Males)**

| | 0 | I | B | IB | C | CI | CB | CIB |
|---|---|---|---|---|---|---|---|---|
| **0** | 1.00 | **** | **** | **** | **** | **** | **** | **** |
| **I** | **** | 1.00 | **** | **** | **** | **** | **** | **** |
| **B** | **** | **** | 1.00 | **** | **** | **** | **** | **** |
| **IB** | **** | **** | **** | 1.00 | **** | **** | **** | **** |
| **C** | **** | **** | **** | **** | 1.00 | **** | **** | **** |
| **CI** | **** | **** | **** | **** | **** | 1.00 | **** | **** |
| **CB** | **** | **** | **** | **** | **** | **** | 1.00 | **** |
| **CIB** | **** | **** | **** | **** | **** | **** | **** | 1.00 |

**Age 50-59 (Females)**

| | 0 | I | B | IB | C | CI | CB | CIB |
|---|---|---|---|---|---|---|---|---|
| **0** | 1.00 | **** | **** | **** | **** | **** | **** | **** |
| **I** | **** | 1.00 | **** | **** | **** | *** | **** | **** |
| **B** | **** | **** | 1.00 | **** | **** | **** | **** | **** |
| **IB** | **** | **** | **** | 1.00 | **** | **** | **** | **** |
| **C** | **** | **** | **** | **** | 1.00 | **** | **** | **** |
| **CI** | **** | *** | **** | **** | **** | 1.00 | **** | **** |
| **CB** | **** | **** | **** | **** | **** | **** | 1.00 | **** |
| **CIB** | **** | **** | **** | **** | **** | **** | **** | 1.00 |

**Age 60+ (Males)**

| | 0 | I | B | IB | C | CI | CB | CIB |
|---|---|---|---|---|---|---|---|---|
| **0** | 1.00 | **** | **** | **** | **** | **** | **** | **** |
| **I** | **** | 1.00 | **** | **** | 1.00 | **** | **** | **** |
| **B** | **** | **** | 1.00 | **** | **** | **** | **** | **** |
| **IB** | **** | **** | **** | 1.00 | **** | **** | ** | **** |
| **C** | **** | 1.00 | **** | **** | 1.00 | **** | **** | **** |
| **CI** | **** | **** | **** | **** | **** | 1.00 | **** | **** |
| **CB** | **** | **** | **** | ** | **** | **** | 1.00 | **** |
| **CIB** | **** | **** | **** | **** | **** | **** | **** | 1.00 |

**Age 60+ (Females)**

| | 0 | I | B | IB | C | CI | CB | CIB |
|---|---|---|---|---|---|---|---|---|
| **0** | 1.00 | **** | **** | **** | **** | **** | ** | **** |
| **I** | **** | 1.00 | **** | **** | **** | **** | **** | **** |
| **B** | **** | **** | 1.00 | **** | 1.00 | **** | **** | **** |
| **IB** | **** | **** | **** | 1.00 | **** | **** | **** | **** |
| **C** | **** | **** | 1.00 | **** | 1.00 | **** | **** | **** |
| **CI** | **** | **** | **** | **** | **** | 1.00 | **** | **** |
| **CB** | ** | **** | **** | **** | **** | **** | 1.00 | **** |
| **CIB** | **** | **** | **** | **** | **** | **** | **** | 1.00 |

Table S4: Results of the Dunn's post-hoc test for pairwise comparisons between models on the RMSE metric for data disaggregated by age and gender. The table is organized by males (left column) and females (right column). P-values are adjusted using the Bonferroni correction. Significance levels are indicated by: ****: $p \leq 10^{-4}$; ***: $10^{-4} < p \leq 10^{-3}$; **: $10^{-3} < p \leq 10^{-2}$; *: $10^{-2} < p \leq 0.05$. P-values greater than 0.05 are shown rounded to two decimal places.

#### S3 Different evaluation metric

While in the main text we have evaluated model performances using the root mean squared error (RMSE) metric, in this section we test the robustness of our results by using a different evaluation metric: the mean absolute error (MAE). The following subsections present a comparison of the two metrics computed on weekly death time series aggregated and disaggregated by gender, by age, and by both gender and age, with comments on eventual discrepancies in the results.

##### S3.1 Models' comparison with aggregated data

The model performance ranking remains consistent across both metrics. This indicates that the error distribution is stable across models, and no specific model is benefiting from having fewer large outliers compared to others.

| | **0** | **I** | **B** | **IB** | **C** | **CI** | **CB** | **CIB** |
|---|---|---|---|---|---|---|---|---|
| MAE | 482.15 | 483.65 | 484.54 | 490.17 | 370.14 | 370.57 | 377.73 | 370.80 |
| RMSE | 536.32 | 537.51 | 537.90 | 545.43 | 434.97 | 439.06 | 450.50 | 441.14 |

Table S5: Performance comparison of the eight model configurations on the aggregated data. The table reports the Mean Absolute Error (MAE) and Root Mean Square Error (RMSE) for the total weekly deaths. Lower values indicate better model performance.

##### S3.2 Models' comparison with gender-disaggregated data

Similarly to the analysis on the aggregated data, the ranking of model performances shown in Table S6 remain consistent across the MAE and RMSE metric when data are disaggregated by gender. There is only a minor difference for models for females: model CB with gender-stratified contacts and behaviors ranks 3rd for females in MAE but 4th in RMSE. Conversely, model I with only gender-stratified IFR ranks 4th in MAE but 3rd in RMSE. This discrepancy indicates that model CB has a lower average error but suffers from larger outliers (which RMSE penalizes heavily) compared to model I, which is more stable with fewer extreme errors.

**Males**

| | **0** | **I** | **B** | **IB** | **C** | **CI** | **CB** | **CIB** |
|---|---|---|---|---|---|---|---|---|
| MAE | 446.22 | 288.17 | 371.19 | 255.55 | 323.25 | 221.61 | 268.27 | 237.25 |
| RMSE | 526.95 | 347.42 | 445.85 | 304.04 | 395.98 | 271.84 | 329.87 | 281.97 |

**Females**

| | **0** | **I** | **B** | **IB** | **C** | **CI** | **CB** | **CIB** |
|---|---|---|---|---|---|---|---|---|
| MAE | 223.28 | 196.37 | 167.64 | 250.89 | 254.69 | 167.22 | 171.92 | 211.32 |
| RMSE | 264.96 | 215.74 | 211.23 | 273.70 | 303.18 | 192.79 | 225.40 | 231.44 |

Table S6: Performance comparison of the eight model configurations on the disaggregated data by gender. The table reports the Mean Absolute Error (MAE) and Root Mean Square Error (RMSE) for male (top) and female (bottom) weekly deaths. Lower values indicate better model performance.

#### S3.3 Models' comparison with age-disaggregated data

Table S7 presents the MAE and RMSE results for the age-disaggregated data. The discrepancies in model ranking between the two metrics are minimal. For the 30–39 age group, the rankings are identical. In other age groups, we observe minor shifts where models exchange rank positions depending on whether MAE or RMSE is used. However, the statistical analysis reveals that the performance differences between these rank-switching models are not statistically significant. Consequently, these variations in ranking should be interpreted as noise rather than meaningful differences in model quality. This is most evident in the 60+ age group: while Model C achieves the lowest RMSE, it ranks third in MAE (behind models CIB and CI). Since the statistical tests confirm that the differences among these three models are non-significant for both metrics, we conclude that models C, CI, and CIB effectively share the top tier of performance for reproducing 60+ mortality data.

**Age group 20-29**

| | **0** | **I** | **B** | **IB** | **C** | **CI** | **CB** | **CIB** |
|---|---|---|---|---|---|---|---|---|
| MAE | 10.55 | 10.46 | 10.60 | 10.60 | 8.92 | 8.86 | 9.11 | 9.07 |
| RMSE | 11.31 | 11.24 | 11.38 | 11.36 | 9.53 | 9.46 | 9.72 | 9.69 |

**Age group 30-39**

| | **0** | **I** | **B** | **IB** | **C** | **CI** | **CB** | **CIB** |
|---|---|---|---|---|---|---|---|---|
| MAE | 25.07 | 24.92 | 25.21 | 25.18 | 22.93 | 22.80 | 23.32 | 23.25 |
| RMSE | 27.12 | 26.99 | 27.31 | 27.20 | 24.59 | 24.41 | 24.93 | 24.91 |

**Age group 40-49**

| | **0** | **I** | **B** | **IB** | **C** | **CI** | **CB** | **CIB** |
|---|---|---|---|---|---|---|---|---|
| MAE | 42.87 | 42.47 | 43.00 | 43.03 | 28.66 | 28.45 | 29.30 | 29.27 |
| RMSE | 46.86 | 46.53 | 47.09 | 46.95 | 31.34 | 31.05 | 31.91 | 31.93 |

**Age group 50-59**

| | **0** | **I** | **B** | **IB** | **C** | **CI** | **CB** | **CIB** |
|---|---|---|---|---|---|---|---|---|
| MAE | 206.75 | 205.05 | 207.09 | 207.03 | 185.50 | 179.98 | 185.86 | 180.95 |
| RMSE | 222.05 | 220.69 | 222.75 | 222.06 | 200.13 | 193.54 | 199.84 | 194.51 |

**Age group 60+**

| | **0** | **I** | **B** | **IB** | **C** | **CI** | **CB** | **CIB** |
|---|---|---|---|---|---|---|---|---|
| MAE | 628.41 | 634.36 | 634.40 | 640.07 | 465.95 | 464.71 | 475.06 | 463.59 |
| RMSE | 726.69 | 733.32 | 732.62 | 740.26 | 521.44 | 523.74 | 536.60 | 522.76 |

Table S7: Performance comparison of the eight model configurations on the data disaggregated by age. Each table reports the Mean Absolute Error (MAE) and Root Mean Square Error (RMSE) for a specific age bracket. Lower values indicate better model performance.

##### S3.4 Models' comparison with age- and gender-disaggregated data

Finally, Table S8 presents the MAE and RMSE results for the age- and gender-disaggregated data. The ranking of model performance is largely robust to the choice of metric. For males in the age groups 40–49, 50–59, and 60+, the rankings are identical. In the younger male groups (20–29 and 30–39), rank discrepancies occur only between models with statistically indistinguishable performances; these models should, therefore, be considered equivalent. Similarly, for females, the rankings are identical across all age groups except 60+. In this oldest cohort, the ranking is unstable: Model C ranks first in MAE but falls to third in RMSE. This suggests that while Model C minimizes error on average for older females, it is more susceptible to large outliers compared to Model 0, which offers a more stable, albeit slightly less precise, fit.

**Age 20-29 (Males)**

| | 0 | I | B | IB | C | CI | CB | CIB |
|---|---|---|---|---|---|---|---|---|
| MAE | 5.41 | 6.36 | 5.82 | 6.81 | 4.58 | 5.41 | 5.06 | 5.89 |
| RMSE | 5.79 | 6.82 | 6.24 | 7.28 | 4.90 | 5.78 | 5.41 | 6.31 |

**Age 30-39 (Males)**

| | 0 | I | B | IB | C | CI | CB | CIB |
|---|---|---|---|---|---|---|---|---|
| MAE | 12.35 | 14.77 | 13.40 | 15.85 | 11.32 | 13.58 | 12.53 | 14.77 |
| RMSE | 13.51 | 16.12 | 14.67 | 17.24 | 12.33 | 14.70 | 13.57 | 15.99 |

**Age 40-49 (Males)**

| | 0 | I | B | IB | C | CI | CB | CIB |
|---|---|---|---|---|---|---|---|---|
| MAE | 17.77 | 22.67 | 19.83 | 24.92 | 11.49 | 15.16 | 13.41 | 17.25 |
| RMSE | 19.31 | 24.80 | 21.68 | 27.18 | 12.44 | 16.45 | 14.48 | 18.68 |

**Age 50-59 (Males)**

| | 0 | I | B | IB | C | CI | CB | CIB |
|---|---|---|---|---|---|---|---|---|
| MAE | 81.02 | 104.58 | 90.93 | 114.96 | 58.54 | 78.44 | 67.71 | 87.60 |
| RMSE | 87.45 | 112.77 | 98.18 | 123.48 | 63.55 | 84.24 | 72.86 | 93.89 |

**Age 60+ (Males)**

| | 0 | I | B | IB | C | CI | CB | CIB |
|---|---|---|---|---|---|---|---|---|
| MAE | 541.01 | 374.69 | 473.23 | 323.87 | 384.47 | 236.28 | 315.18 | 211.66 |
| RMSE | 627.59 | 449.70 | 552.43 | 387.70 | 455.82 | 293.07 | 384.40 | 258.19 |

**Age 20-29 (Females)**

| | 0 | I | B | IB | C | CI | CB | CIB |
|---|---|---|---|---|---|---|---|---|
| MAE | 5.14 | 4.11 | 4.77 | 3.80 | 4.34 | 3.45 | 4.05 | 3.18 |
| RMSE | 5.60 | 4.52 | 5.21 | 4.18 | 4.73 | 3.81 | 4.42 | 3.54 |

**Age 30-39 (Females)**

| | 0 | I | B | IB | C | CI | CB | CIB |
|---|---|---|---|---|---|---|---|---|
| MAE | 12.72 | 10.15 | 11.80 | 9.33 | 11.60 | 9.21 | 10.79 | 8.47 |
| RMSE | 13.74 | 11.02 | 12.78 | 10.12 | 12.40 | 9.87 | 11.51 | 9.10 |

**Age 40-49 (Females)**

| | 0 | I | B | IB | C | CI | CB | CIB |
|---|---|---|---|---|---|---|---|---|
| MAE | 25.94 | 20.29 | 23.82 | 18.47 | 18.33 | 13.94 | 16.82 | 12.57 |
| RMSE | 28.06 | 22.07 | 25.82 | 20.07 | 19.72 | 15.12 | 18.08 | 13.73 |

**Age 50-59 (Females)**

| | 0 | I | B | IB | C | CI | CB | CIB |
|---|---|---|---|---|---|---|---|---|
| MAE | 125.73 | 100.47 | 116.16 | 92.07 | 126.96 | 101.54 | 118.15 | 93.35 |
| RMSE | 135.26 | 108.34 | 125.10 | 98.94 | 137.53 | 109.91 | 127.78 | 101.17 |

**Age 60+ (Females)**

| | 0 | I | B | IB | C | CI | CB | CIB |
|---|---|---|---|---|---|---|---|---|
| MAE | 166.13 | 275.16 | 202.36 | 341.03 | 159.40 | 232.43 | 173.09 | 294.72 |
| RMSE | 194.95 | 298.80 | 219.92 | 371.73 | 206.66 | 257.43 | 198.01 | 322.87 |

Table S8: Performance comparison of the eight model configurations on data disaggregated by both age and gender. The table is organized by males (left column) and females (right column) across five age groups. Reported metrics are Mean Absolute Error (MAE) and Root Mean Square Error (RMSE). Lower values indicate better model performance.
